# Intervention on Modifiable Lifestyle and Physiological Factors via Variational Autoencoder Reveals Changes in Functional Connectivity-Mediated Risk for Alzheimer’s Disease

**DOI:** 10.1101/2025.09.16.25335925

**Authors:** Anton Orlichenko, Shengxian Ding, Emily Johns, Zhiling Gu, Xinyuan Tian, Xiaoxuan Li, Yize Zhao

## Abstract

Alzheimer’s disease (AD) remains without effective treatment, largely due to the fact that clinical symptoms emerge only after decades of silent pathological progression. It is urgently needed to identify modifiable risk factors in earlier life stages, when preventive interventions may still be effective. Functional connectivity (FC) has emerged as a promising neuromarker for both neurodegenerative processes and behavioral traits, making it a potential bridge between early-life health profiles and late-life AD risk. In this work, we introduce a novel integrative framework that models how early-life lifestyle and physiological factors influence AD risk through their impact on brain FC. Our approach combines a modified variational autoencoder (VAE) that simulates FC changes under interventions with a predictive model that estimates AD risk based on FC patterns. This design enables training of the generative and predictive components under different datasets and populations, with FC acting as the bridge between early-life modifiable factors and late-life disease risk. Applying our framework to data from the Human Connectome Project (HCP), UK Biobank (UKB), and Alzheimer’s Disease Neuroimaging Initiative (ADNI), we validate its ability to capture known risk factors, such as age and polygenic risk score, on FC-mediated AD risk. We also identify earlier-life modifiable factors including tobacco use, sleep quality, physical activity and weight/BMI that significantly influence AD risk. Notably, we observe a U-shaped relationship between blood pressure and AD risk, and highlight the brain visual and somatomotor networks as key mediators of risk through FC. Our approach provides a powerful tool for investigating the effect pathways linking early-life interventions to neurodegenerative outcomes, with broad applicability to other brain-related disorders.

## 1 Introduction

Alzheimer’s disease (AD) is a progressive neurodegenerative disease characterized by cognitive decline, loss of memory, and impaired reasoning skills (Safiri et al., 2024). The pathological hallmarks of AD are widely recognized to include the accumulation of amyloid-*β* and tau peptide (Y. Chen & Yu, 2023; M. P. Murphy & LeVine, 2010). An estimated 6.5 million Americans are currently living with AD, imposing substantial economic burdens that amount to hundreds of billions of dollars annually. Existing treatments, such as acetylcholinesterase inhibitors, non-steroidal anti-inflammatory compounds, sex steroid hormones, and antioxidants (Allain et al., 2007), demonstrate limited efficacy in altering the progression of AD and mild cognitive impairment (MCI). Moreover, AD symptoms typically manifest many years after the initial onset of pathological changes, meaning that therapeutic interventions often occur too late to be fully effective (Thangwaritorn et al., 2024). Consequently, there has been significant research interest in developing effective interventions during early-life stages, before clinical symptoms manifest (Gandy et al., 2017; Seifan et al., 2015). Early interventions well before the onset of noticeable cognitive symptoms can be essential for altering disease outcomes timely.

The stages of AD progression include the preclinical, prodromal, and patent dementia periods (Vermunt et al., 2019). The preclinical AD stage can begin as early as midlife and span decades without overt symptoms, despite accumulating pathological features such as amyloid plaques and neurofibrillary tangles in the brain (Thal et al., 2013). The prodromal stage involves MCI, where measurable deficits are present but daily independence is maintained (Albert et al., 2011), followed by dementia, characterized by severe impairments in cognition and everyday functioning. The long, symptom-free preclinical stage (averaging 10 years) poses a major challenge to early detection. Most studies attempting to capture this stage must enroll large cohorts of cognitively normal individuals, of whom only about 10% will eventually develop AD (Bennett et al., 2018; Dhana et al., 2023; Petersen et al., 2010). This makes such research inefficient and resource-intensive. As a result, there is growing interest in using biomarkers to more precisely identify individuals at risk during these early stages (Millar et al., 2022; Risacher & Saykin, 2013).

Among these biomarkers, neuroimaging including functional MRI (fMRI) has shown great promise to reveal altered activation patterns in memory-related brain regions among individuals at risk (Noh et al., 2023). Functional connectivity (FC), an fMRI-based imaging feature derived to characterize network-level temporal synchronization of activity, has emerged as a particularly powerful neuromarker. FC consistently reveals network-level disruptions during the transition from cognitively normal aging to MCI (Penalba-Sánchez et al., 2022), and has demonstrated strong predictive accuracy in identifying individuals likely to progress from prodromal AD to dementia (Noh et al., 2023). Beyond AD, FC serves as a robust individual fingerprint, capturing both normative traits such as age (Hu et al., 2019), sex (İçer et al., 2020), race (Orlichenko et al., 2023), and cognitive ability (Qu et al., 2024), as well as deviations seen in disorders like ADHD (Castellanos & Aoki, 2016), schizophrenia (Collin et al., 2020), and ASD (Sung & Park, 2024). Given the sensitivity of FC to both normal variation and disease states, and its quantifiable progression across AD stages, we propose using FC as a bridge to link early-life behaviors and physiological traits to later AD risk. Critically, most current AD studies focus on older adults who are already on the disease trajectory (Bennett et al., 2018; Petersen et al., 2010), limiting our understanding of how modifiable early-life factors shape long-term vulnerability.

In this study, we leverage FC as an intermediate neurobiological representation to bridge the potential causal pathway from early-life behaviors and physiological measurements to late-life AD risk. By rigorously modeling how modifiable factors shape FC and how FC influences AD vulnerability across studies over lifespan, we aim to elucidate causal mechanisms by which early interventions may alter long-term disease outcomes. As illustrated in Figure 1, we develop a behavior- and lifestyle factor-conditioned variational autoencoder (VAE) to model how modifiable factors influence FC and, in turn, AD risk. In the current literature, such kind of analytical objective has mainly achieved by Mendelian randomization (MR) as discussed in Section 2, which tends to be constrained by the availability of strong genetic instrumental variables for diverse behaviors or lifestyle factors (Korologou-Linden et al., 2022; Sanderson et al., 2022), known as the weak IV problem (Andrews et al., 2019). This limitation has hindered efforts to establish causal links between early-life exposures and late-life clinical outcomes in general. In contrast, behaviors and lifestyle factors are strongly correlated with FC endophenotype, and a growing body of evidence supports the robust prediction of FC for AD. Our approach integrates these insights by modeling how behavioral interventions affect FC and subsequently influence AD risk. Specifically, we train a VAE to generate FC conditioned on behavioral inputs, then use an AD prediction model to quantify changes in risk based on the altered FC. This framework allows us to infer how modifying early-life factors could causally influence AD susceptibility through their effect on brain connectivity.

**Figure 1:**
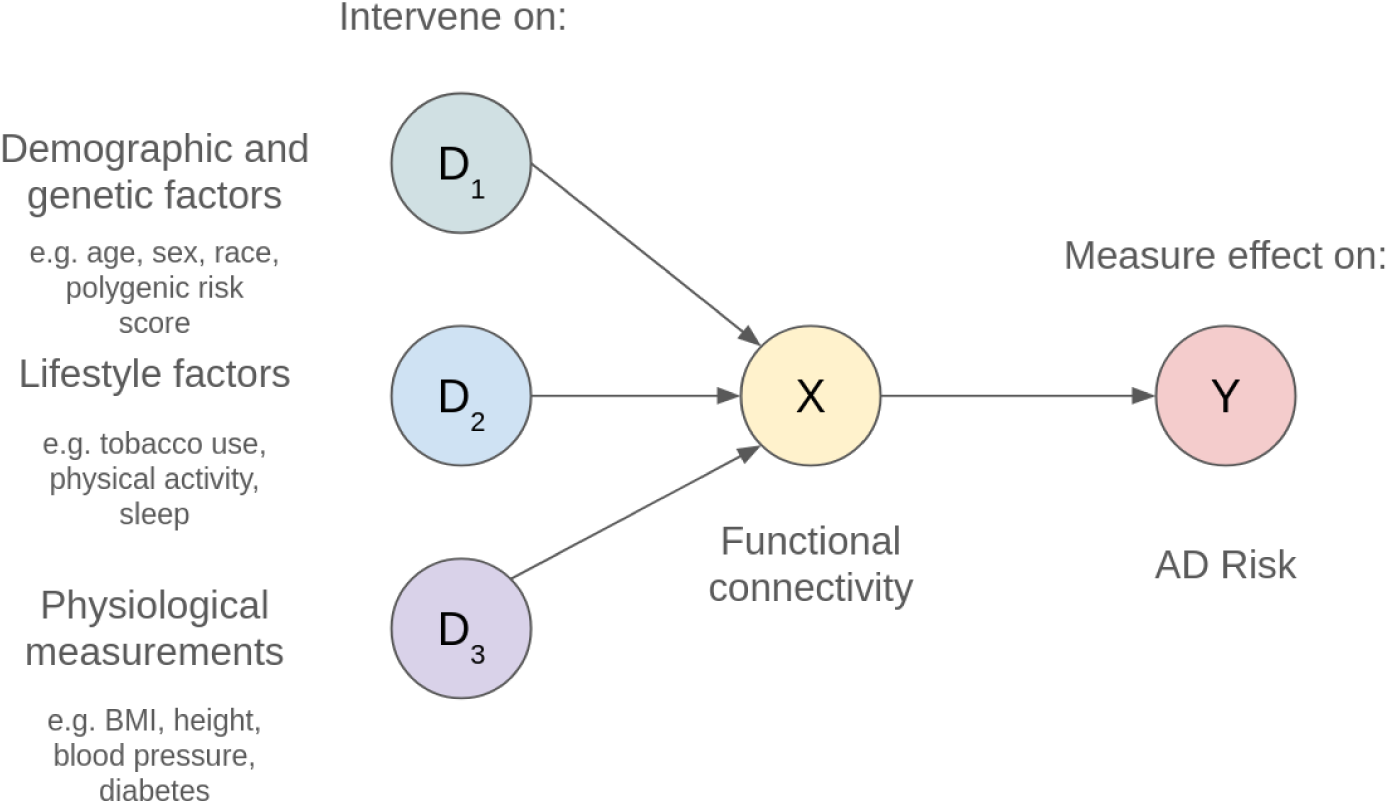
High level overview of the causal effects in our model. We assume that demographic, genetic, and lifestyle factors *D* all have an effect on FC *X*. Additionally, FC is assumed to be representative of cognitive changes occurring in AD represented by risk *Y*. By creating a model that is able to synthesize FC dependent on *D*, we can quantify the increased or decreased risk of AD based on disentangled interventions on *D*. The direct effect of *D* on *Y* is unobserved.

The contributions of our work are twofold. First, although VAEs have previously been applied in causal inference settings (Louizos et al., 2017; Yang et al., 2021), to the best of our knowledge, this is the first study to use a VAE for simulating counterfactual phenotypes in silico–to estimate how hypothetical changes in exposure variables influence the phenotypes, providing a flexible and generalizable generative framework for interrogating complex variable-phenotype relationships. Second, this is the first framework to explicitly model how early-life behavioral and physiological factors influence late-life Alzheimer’s disease risk mediated by brain functional connectivity, enabling quantitative estimation of how early interventions may modulate disease vulnerability. While we focus on Alzheimer’s disease and functional connectivity in this study, the framework is generalizable to other neurological conditions and neuromarker modalities.

The remainder of this paper is organized as follows. Section 2 reviews related work on lifespan data integration. Section 3 describes the interventional VAE framework, FC-mediated AD risk model, and the datasets used, including the Human Connectome Project (HCP), UK Biobank (UKB), and Alzheimer’s Disease Neuroimaging Initiative (ADNI). Section 4 presents the results of our analyses, and Section 5 concludes with discussion and potential future extensions.

## 2 Related Work

Integrating early-life behaviors with disease risks that manifest only in old age presents a significant challenge, though it has been explored in prior work. One widely used method is MR, which uses genetic variants as instrumental variables (IVs) to proxy for exposures, offering an alternative to randomized controlled trials (Sanderson et al., 2022). MR has been used in numerous applications, including AD (Korologou-Linden et al., 2022; Su et al., 2023), schizophrenia (Veeneman et al., 2022), and cancer (Wade et al., 2022). For example, (European Alzheimer’s & Dementia Biobank Mendelian Randomization (EADB-MR) Collaboration et al., 2023) employed MR to assess the influence of 12 modifiable lifestyle factors such as lipid levels, cholesterol, alcohol consumption, and smoking on AD risk. Most of these factors did not show statistically significant associations with AD risk, except for High Density Lipoprotein and ApoE genotype. We attribute this, in part, to the weak instrument problem: it is often difficult to identify genetic variants that are strongly associated with complex behaviors like smoking (Andrews et al., 2019). Additionally, issues such as pleiotropy and violations of the gene-environment equivalence assumption further limit the reliability of MR for certain lifestyle exposures (Sanderson et al., 2022; VanderWeele et al., 2014). In contrast, our current study focuses only on behavioral and lifestyle factors that show significant correlations with brain FC (see Supplementary Materials). Behaviors that do not explain variance in FC are effectively neutralized during inference, with their influence on downstream outcomes averaging to near zero.

Another important line of related research involves mediation analysis, which typically decomposes the impact from an exposure to an outcome into direct and indirect components via a mediator (Rijnhart et al., 2021). Previous studies have used FC between specific brain regions as mediators for cognitive ability in MCI (Huang et al., 2024). More recently, high-dimensional mediation analysis techniques, including those based on Bayesian methods and deep learning, have enabled the use of entire FC networks as mediators (Nath et al., 2023; Tian et al., 2024). Some studies have also examined how FC mediates the relationship between genetic factors and disease outcomes (Dai & Zhang, 2024). However, a critical limitation of classical mediation analysis in the context of our research is that it requires all three components, i.e. exposure, mediator and outcome, to be measured within the same individual. This requirement is impractical when studying how early-life behaviors impact late-life outcomes like AD, due to the decades-long temporal gap and data availability constraints. In contrast, our method overcomes this constraint by leveraging separately collected population datasets under different life stages, deriving generative modeling to infer plausible mediating trajectories and estimate causal effects across lifespan-separated variables.

A core part of our method is VAE. The VAE is a deep generative latent-variable model that learns a probability distribution from which it can sample new data points (Kingma & Welling, 2019). VAEs have been widely adopted in medical research for purposes such as generating synthetic images for data augmentation (Rais et al., 2024), dimensionality reduction, and learning interpretable latent spaces (Doncevic & Herrmann, 2023; G. Kim & Chun, 2023). Notably, VAEs can also model causal effects by accounting for unobserved confounders (H. Kim et al., 2021; Louizos et al., 2017). This property is advantageous in our interventional VAE model: covariates not explicitly conditioned on are nonetheless captured in the latent distribution, mitigating confounding bias.

Finally, a substantial body of research exists on the integration of multimodal data, such as multi-omics (Ballard et al., 2024; Wang et al., 2021; Zheng et al., 2024), or imaging with omics data (Paverd et al., 2024; Watson et al., 2021), including in the context of fMRI and brain-related gene expression in AD studies (Chan et al., 2022; Povala et al., 2024; Yoon et al., 2024). Other studies have combined multiple imaging modalities to improve predictive performance (Qu et al., 2025). However, these approaches primarily integrate data “horizontally” by combining subjects from different datasets into a larger aggregated dataset, which is fundamentally different from our objective of linking subjects from different databases measured under various life stages “vertically” to mimic different information collection at a subject-level. Our approach offers a novel method for integrating temporally and biologically disparate data into a coherent, predictive framework.

## 3 Materials and Methods

This section describes the structure of the lifestyle factor-conditioned VAE for generation of FC data, the datasets used to train the VAE, the dataset used for the AD risk model, and the experiments to intervene on modifiable lifestyle factors and quantify the resulting change in AD risk. We first outline the structure of the VAE used for intervention analysis. We then describe three datasets used for our investigation of early-life risk factors for AD. The first of these is the ADNI dataset of AD patients as well as age-matched cognitively normal controls, used to create our FC-based model of AD. The second dataset is the UKB cohort of older but still largely cognitively normal adults, containing both neuroimaging data and a large dataset of behaviors and physiological risk factors for each subject. The last one is the HCP dataset of young adults with additional behavioral and physiological risk factors, used to identify AD risk in an early-life stage. Preliminary simulation experiments to test our model’s ability to identify causal changes in risk in the presence of a known ground truth are given in the Supplementary Materials.

### 3.1 Variational Autoencoder for Intervention Analysis

An overview of our proposed use of a VAE for AD risk quantification is shown in Figure 2. The idea is to condition a VAE on, in our case, non-modifiable/genetic factors, modifiable risk factors, and physiological measurements in order to synthetically generate FC consistent with intervention on those factors. By training a generative model, we can vary one particular risk factor while holding all others constant, thereby disentangling the effects of each modifiable or non-modifiable risk factor.

**Figure 2:**
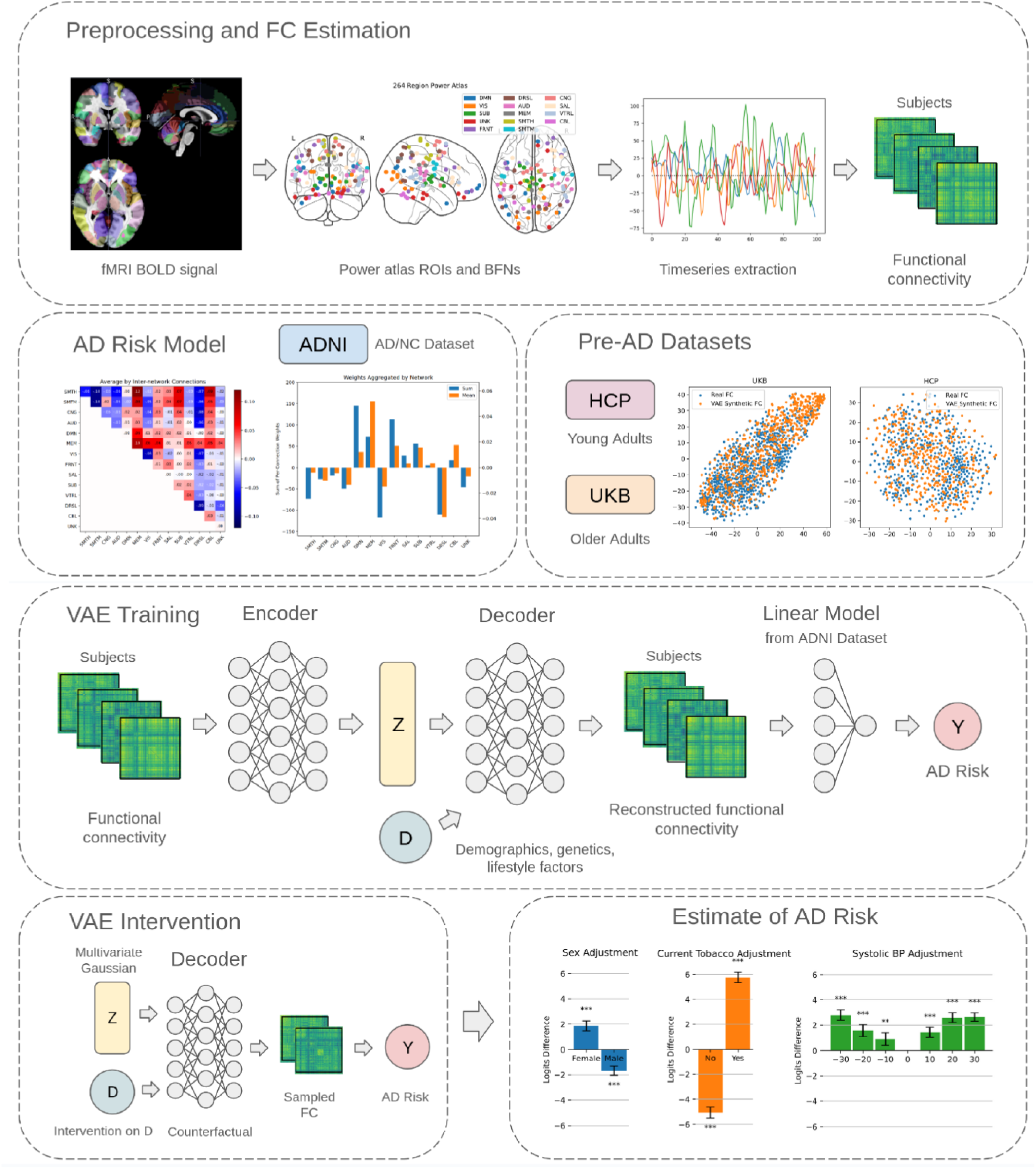
An overview of our proposed VAE-based model for intervention on demographic, genetic, and early-life lifestyle factors *D* to quantify AD risk mediated by FC. An AD risk model is created using the ADNI dataset and risk is evaluated on the HCP and UKB datasets. A VAE consisting of an encoder and decoder reconstructs FC from a latent space that has a target distribution of a multivariate Gaussian. The decoder is conditioned upon factors *D* and the latent state *Z* is made to be uncorrelated with *D* during training. During counterfactual intervention, the change in AD risk is measured for a particular intervention on *D*.

#### 3.1.1 Structure of the Risk Factor-Conditioned VAE

An autoencoder (AE) converts raw features into a lower-dimensional latent space via a learned encoder function **z** = *E_ϕ_*(**x**), along with a decoder function to convert the latent features back into a reconstructed version of the input **x̂** = *D_θ_*(**z**). The AE is trained to minimize the difference between the reconstruction **x̂** and original input **x**, where in our study **x** is the vectorized subject FC. By contrast, a VAE trains the encoder function *E_ϕ_*(**x**) to produce latent features that approximate a known probability distribution *p_θ_*(**z**), most often taken to be a standard multivariate Gaussian distribution *p_θ_*(**z**) = 𝒩 (**0**, **I**) (Kingma & Welling, 2014). This allows for artificially constructing latent samples **z**_samp_ from the approximated distribution, followed by the conversion of those latent features to samples of the original distribution *p_θ_*(**x**|**z**) by passing through the decoder function **x**_samp_ = *D_θ_*(**z**_samp_). A VAE is trained by maximizing the Evidence Lower Bound (ELBO). Maximizing the ELBO is equivalent to maximizing the reconstruction probability log*p_θ_*(*x*|*z*) while minimizing the KL divergence between our empirical and target distributions, most often the standard Gaussian distribution mentioned previously.

While prior research has explored conditioning VAE models on user-specified inputs (Razavi et al., 2019; Sohn et al., 2015), and practical applications have demonstrated zero-shot generation capabilities (Ramesh et al., 2021), these approaches typically lack explicit consideration of subject-specific phenotypes. Similarly, VAEs have been employed to generate synthetic FC data for data augmentation (J.-H. Kim et al., 2021), but often without integrating phenotype information. In contrast, our model introduces a novel phenotype-aware conditioning mechanism, enabling the generation of synthetic FC data that more accurately reflects individual variability and improves downstream analysis.

In this work, we include phenotypic features as input to the decoder function **x̂** = *D_θ_*(**z**, **d**), where **z** are the latent features and **d** are the subject phenotypes. During training, we decorrelate the latent state **z** = *E_ϕ_*(**x**) from phenotypes features **d** so that all of the FC signal that can be attributed to phenotypes is based on user-provided input and not on the encoded latent features. To this end, we make several modifications to the traditional VAE loss function.

#### 3.1.2 Incorporate Phenotype Information

First, the term representing reconstruction error in the loss function remains unchanged with regard to the ELBO formulation, except for the introduction of phenotype information:

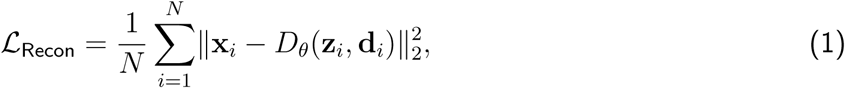

here *N* is the number of subjects, **x***_i_* are the input features (vectorized FC), **z***_i_* = *E_ϕ_*(**x***_i_*) are the latent features derived from the encoder, and **d***_i_* are subject *i*’s phenotypes.

#### 3.1.3 Extension to Multidimensional Latent Space

Second, when considering a multivariate standard normal distribution *p_θ_*(**z**) = 𝒩 (**0**, **I**) for the latent features, the KL divergence part takes the form (K. P. Murphy, 2023):

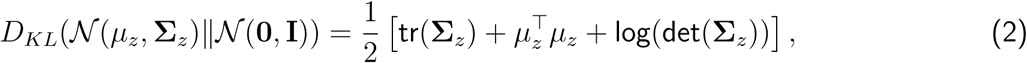

which is challenging because it requires computing and backpropagating through the log-determinant of the empirical latent covariance matrix **Σ***_z_*.

We note that the ELBO loss function of the standard VAE is applicable to scalar latent features *z* and not multi-dimensional latent features 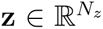. This may allow for a non-diagonal covariance matrix in the empirical distribution of latent features *q_ϕ_*(**z**|**x**) = 𝒩 (**0**, **Σ**). Thus, we modify a part of the ELBO loss function to specifically target a diagonal covariance matrix and zero expected value for the latent features:

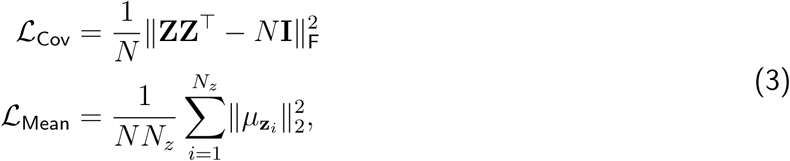

Where 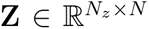 is the Gram matrix of all *N_z_* latent features for all *N* subjects, **z***_i_* is the vector of latent feature *i* for all *N* subjects, and 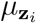 is its mean.

#### 3.1.4 Decorrelate Latent Features from Phenotypes

Third, we introduce a term that penalizes correlations between the empirical latent features and the modifiable, non-modifiable, or physiological measurement features used to condition the VAE. We define

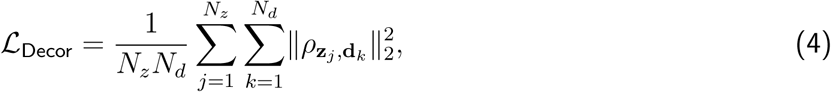

Here 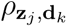 is the empirically calculated correlation between between latent feature **z***_j_* and phenotypic feature **d***_k_* for all *N* subjects.

#### 3.1.5 Classifier Guidance

Finally, during training of the intervention VAE, we generate synthetic samples by randomly selecting phenotypic inputs, and penalize prediction errors relative to pre-trained models. For a single phenotypic prediction derived from a synthetic latent conditioned on user-specified phenotypes, 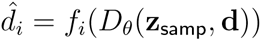, we define

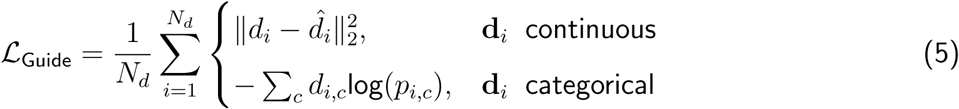

here the functions *f_i_*(·) are models trained on ground-truth subject FC data, *d_i,c_* denotes the one-hot encoded true class label for phenotype *i*, *p_i,c_* represents the predicted probability for class *c* and phenotype *i*, and the loss is defined as Mean Squared Error (MSE) for continuous phenotypes and Cross-Entropy (CE) loss for categorical phenotypes

The final loss function for training the phenotype-conditioned VAE is formulated as:

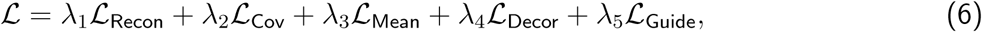

where *λ*_1−5_ are the hyperparameters chosen random grid search alongside learning rate and latent dimension size.

#### 3.1.6 Inference for FC-Mediated AD Risk

Once trained, the VAE model is used to generate synthetic FC based on counterfactual intervention in subject phenotypes. This FC is then evaluated using an AD risk model to determine whether the intervention increased or decreased AD risk. This is modeled by

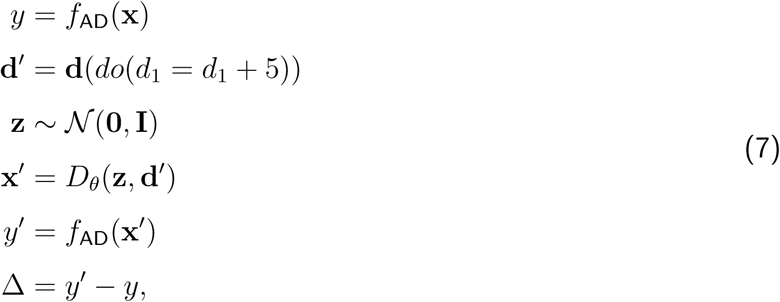

where **d** are the phenotypic factors, **d**^′^ = **d**(*do*(*d*_1_ = *d*_1_ + 5) are the phenotypic factors after intervention or adjustment (the +5 is an just an example adjustment for a continuous variable), **z** is a sample from an isotropic multivariate normal distribution, **x**^′^ is a single subject’s sampled synthetic FC conditioned on **d**^′^, and *y*^′^ is the quantified FC-mediated AD risk for the subject with phenotypic factors **d**^′^, estimated from **x**^′^ using risk model *f*_AD_. This is compared with the original FC-mediated AD risk *y* estimated from the original subject FC **x** to derive the logits change Δ due to the intervention, averaged over the entire cohort.

The VAE model was trained on a Linux workstation with 64GB of RAM and an Nvidia RTX 3060 graphics card. The hyperparameters used for training were *λ*_Recon_ = 1000, *λ*_Cov_ = 10000, *λ*_Mean_ = 1, *λ*_Decor_ = 10, and *λ*_Guide_ = 0.01. Additionally, classifier guidance models used the scikit-learn Ridge regressor with *α* = 100 for continuous variables and scikit-learn LogisticRegression classifier with *C* = 100 for categorical variables (Pedregosa et al., 2011). Hyperparameters were chosen using random grid search based on how well the synthetic samples created using the VAE recapitulated group differences in the original FC data.

### 3.2 ADNI Dataset and Risk Predictive Model

The ADNI (adni.loni.usc.edu) is an ongoing study with primary goal to determine whether clinical, imaging, genetic and biochemical biomarkers can be combined to understand the pathology of AD and track the progression of the disease at an early stage. In our current study, we focus on subjects with fMRI scans acquired, and 699 subjects were included with an average age of 77.5 ± 11.1 years. Scanning was conducted at multiple sites using GE, Philips, and Siemens scanners. The TR was variable and ranged from 6s to 0.6s depending on scanner, subject, and date of scan. Coregistration and warping to MNI space of resting state fMRI scans was carried out using SPM12 (Friston, 2003). BOLD signal was extracted using the 264 ROI Power atlas (Power et al., 2011), and the resulting timeseries were bandpass filtered between 0.01 and 0.15 Hz or the highest frequency available in the data to eliminate scanner drift, noise, heartbeat, and breathing signal. Pearson correlation between pairs of ROIs were taken to create per-subject FC maps.

A total of 737 scans consisting of scans of subjects with current AD diagnosis or current cognitively normal status were processed. Scans of subjects corresponding to mild cognitive impairment were excluded. These scans were divided into 87 AD male, 68 AD female, 235 cognitively normal male, and 347 cognitively normal female scans. In order to avoid the confounding effect of sex on AD diagnosis present in this dataset (relatively more AD males compared to AD females, and relatively more cognitively normal AD females compared to males), a balanced dataset was created with randomly selected 68 subject scans from each category, for a total of 272 scans. A linear logistic regression model was then trained using the scikit-learn package (Pedregosa et al., 2011) to predict AD from the scans of these 272 subjects. Subjects were then randomized and the learning process repeated for 20 bootstrap repetitions. Finally, AD-predicting FC model weights from all logistic regression models were averaged to create a de-biased model that was representative of the whole subset of the ADNI dataset used in this work.

### 3.3 UKB Dataset

The UKB is a large-scale biomedical database from over 500,000 participants, primarily collected during mid-to-late adulthood. Resting state fMRI data as well as demographics, lifestyle factors, and physiological measurements was collected for 40,623 UKB participants. A fraction of UKB subjects had both a first and second fMRI scan, but only a single scan, the first, was used to train the VAE model. Correlation between 124 fields, consisting of questionnaire fields, demographics, and physiological measurements, was performed with resting state fMRI-derived FC. A set of 20 fields significantly correlated with FC were selected for inclusion in the VAE model and further analysis. These fields and their summary statistics are given in Supplementary Materials Table 6. A table of all correlations including field ID and description included in the preliminary investigation of fields associated with FC change is provided in the Supplementary Materials.

Only those subjects that had both a resting state fMRI scan and values for all fields were retained in the analysis, leading to the inclusion of 29,466 subjects with average age 64.9 ± 7.5 years. UKB resting state fMRI was acquired at a resolution of 2.4 by 2.4 by 2.4 mm with an 88 by 88 by 64 FOV matrix. The duration of the scan was 6 minutes, using a TR of 0.735s, resulting in 490 timepoints per scan. The TE was 39ms, and the scanner protocol used GE-EPI with x8 multisclice acceleration, no iPAT, a flip angle of 52°, and fat saturation (Alfaro-Almagro et al., 2018; Miller et al., 2016; Smith et al., 2022).

Although the UKB provides precomputed FC as data fields, these FC are based on either 25 or 100 component ICA, and are not partitioned into canonical functional networks. Therefore, we preprocessed the original resting state fMRI scan data using SPM12, including scan volume coregistration and warping to MNI space (Friston, 2003). The BOLD signal was then extracted from 264 ROIs defined in the Power atlas (Power et al., 2011) to create timeseries at each ROI. These timeseries were bandpass filtered between 0.01 and 0.15 Hz to remove scanner drift, noise, heartbeat, and signal associated with breathing. The Pearson correlation between each pair of ROIs was then taken to create FC maps for each subject. These FC maps along with the 20 data fields extracted earlier were then used in the following VAE-based intervention analysis. For reference, the brain functional networks making up the Power atlas and the distribution of ROIs therein is given in Supplementary Materials Table 9.

### 3.4 HCP Dataset

In addition to the UKB dataset, we tested our VAE-based intervention in the HCP young adult dataset (Van Essen et al., 2012). This gives insight into AD-associated FC changes in a population before that population is of an age to show clinical signs of AD or MCI. Resting state fMRI as well as physiological measurements were acquired for 456 subjects from the HCP young adult dataset, aged 28.5 ± 3.69 years. The 13 fields selected for inclusion into the VAE-based analysis are listed in Supplementary Materials Table 7. These were chosen in order to correspond most closely to the 20 chosen fields in the UKB dataset listed in Supplementary Materials Table 6, although, since the data collection for HCP was not as thorough as for UKB, not all UKB fields had an analogue in the HCP dataset.

Resting state fMRI for the HCP dataset was acquired using a Siemens Skyra 3T scanner using 3mm isotropic resolution at a TR of 0.7s (Uğurbil et al., 2013). The scan time of 29 minutes resulted in subjects with an fMRI time course of 1200 time points. Temporal data was bandpass filtered between 0.01 and 0.15 Hz to eliminate scanner drift, noise, heartbeat, and breathing signal. Coregistration and warping to MNI space were performed using SPM12 (Friston, 2003). As for the UKB data, BOLD signal was extracted using the 264 ROI Power template (Power et al., 2011), and the Pearson correlation between pairs of ROIs was used to create per subject FC maps. The HCP 500 data release was used for our study, having 493 subjects with available resting state fMRI data. Not all of these subjects had valid values for all phenotypic fields, resulting in a final count of 456 subjects for our downstream analysis. Although the selected fields in the HCP cohort were matched closely to those in the UKB cohort, measurement units often differed (metric in UKB, imperial in HCP) and were retained as originally reported to facilitate comparisons with other studies.

Although we have many more subjects in the UKB dataset than in the HCP dataset, only 700 subjects were used at a time to train the VAE in order to have comparable sizes for the two datasets as well as for speed of convergence. These subjects were chosen randomly from the pool of available subjects each time the VAE was trained to avoid bias. Additionally, the model was only ever conditioned on a maximum of five phenotypes at once. For the UKB dataset, these were always age, sex, waist circumference, and height, with the fifth phenotype changed based on what field was being intervened on. For the HCP dataset, these were age, sex, weight, and height, with the fifth phenotype again being variable. This was undertaken because of slow convergence/instability in training when using the whole suite of phenotypic fields.

## 4 Results

### 4.1 AD Risk Model

The ADNI-based model for FC-mediated AD risk is shown in Figure 3. The weights of each functional connection contributing to AD risk are shown in Figure 3A and the network-averaged weights in Figure 3B and 3C. It can be seen that the SMTH-SMTH, DRSL, and VIS-VIS connections are highly negative for FC-mediated AD risk. Indeed, (N. A. Singh et al., 2023) has found that decreased VIS-VIS connectivity is a robust consequence of AD progression. Meanwhile, most functional connections related to the MEM network are positively correlated with AD risk, although this is less significant since fewer ROIs are part of the MEM network. Also somewhat highly correlated with increased AD risk are DMN connections. Although the mean network weights for DMN-associated connections are less positive than for the MEM network, the high number of DMN ROIs means overall the DMN network contributes the most to positive signal for AD prediction. Also notable is the FRNT network which is second behind the DMN in overall AD risk by sum of connection weights. Supplementary Materials Table 8 shows the top ten negatively and positively weighted functional connections for AD risk.

**Figure 3:**
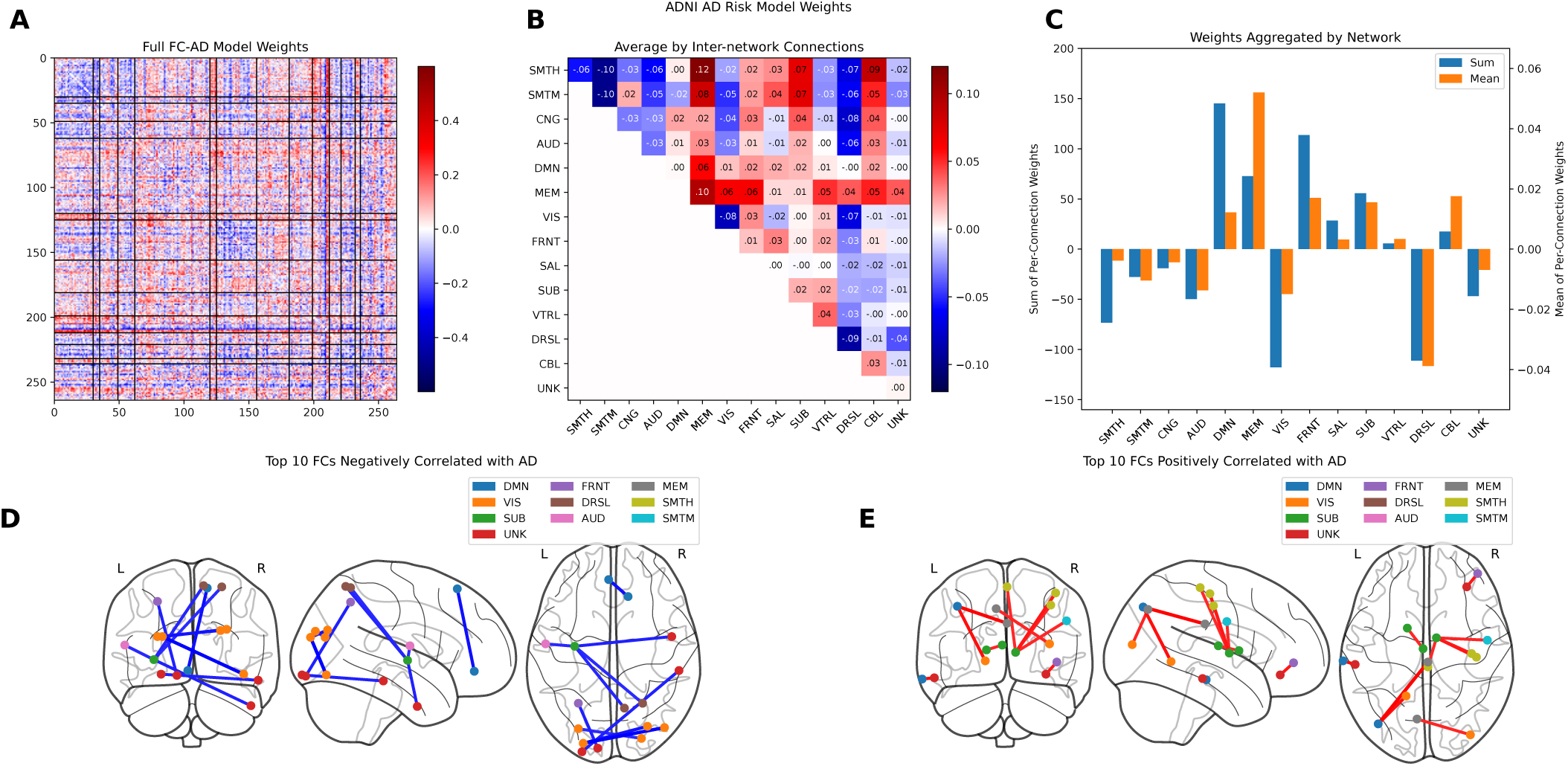
Model weights showing relationship between FC and AD diagnosis in the ADNI dataset (A). These FC-based weights for predicting AD are used to determine AD risk in the VAE-based analysis. The data is broken down by Power functional network in (B). Overall positive or negative contribution by brain functional network is shown in (C). Part (D) shows the top 10 network connections negatively correlated with AD status. Notice that most of the connections are long range and that most ROIs are part of the VIS or UNK networks. Part (E) shows the top 10 network connections most positively correlated with AD status. Note that most connection weights negatively correlated with AD are long range connections, while weights positively correlated with AD tend to be short-range. Inter-VIS and DRSL network connections tend to be have negative weights for predicting AD. Connections involving the MEM network are especially likely to be positively correlated with AD status. Additional details of the top 10 positive and negative connections is given in Supplementary Materials Table 8.

These averaged connections weights are multiplied by subject FC, before or after VAE-based intervention, to find the logits *y* of AD risk, i.e. *y* = **w**^T^**x**, where **w** are the averaged connection weights from the ADNI model and **x** is the subject FC before or after intervention. Since the logits difference in AD risk is relative to the baseline value without intervention, and is dependent on the number of subjects being evaluated, we mention here that the baseline logits score without intervention was −42.6 ± 0.8 per subject for the UKB dataset and −72.9 ± 1.6 per subject for the HCP dataset. Any logit difference reported subsequently is higher or lower than the baseline values reported here. ADNI dataset subjects that were diagnosed with AD had an average logit score of 68.1 ± 47.9 and cognitively normal ADNI subjects had an average score of −52.5 ± 50.0. Note that in case of ADNI the standard error is standard deviation of scores among subjects while in the UKB and HCP dataset the error listed is the variance between bootstrap VAE-sampling repetitions.

### 4.2 Change in AD Risk Following VAE-based Intervention

In this section, we describe the changes in FC-mediated AD risk following intervention on non-modifiable/genetic, modifiable lifestyle, and physiological measurement fields in both the UKB and HCP datasets.

#### 4.2.1 Earlier-life Risk Factors Leveraging UKB Dataset

Figure 4 illustrates how modifiable factors and physiological measures affect AD risk via FC-mediated modeling in the UKB dataset. Tobacco use has one of the strongest effects, with current smoking increasing AD-like FC patterns; past smoking has a smaller but still significant impact, consistent with findings linking heavy smoking to dementia risk (Zhong et al., 2015). We find that UKB-provided polygenic risk scores for AD (Collister et al., 2022) are associated with higher FC-mediated AD risk, as expected.

**Figure 4:**
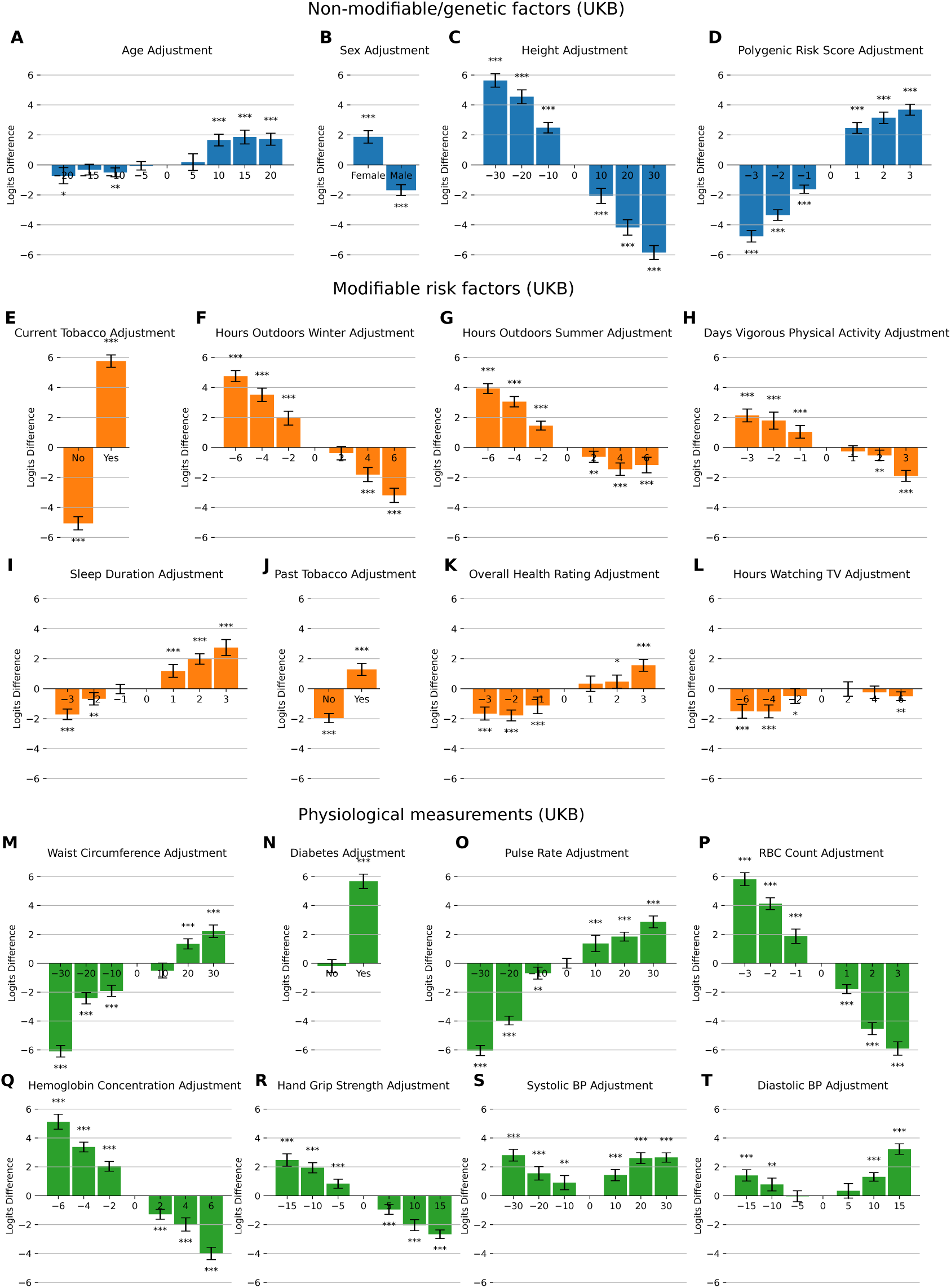
Changes in AD risk as quantified by FC mediator for non-modifiable/genetic factors, modifiable risk factors, and physiological measurements in the UKB dataset. For each measurement, 20 bootstrap repetitions of an intervention, magnitude shown on the horizontal axis, were made and the mean and standard deviation recorded. Error bars represent the standard error, and stars represent significance level: one star (*p <* 0.05), two stars (*p <* 0.01), three stars (*p <* 0.001). Refer to Supplementary Materials Table 6 for summary statistics and units.

Low physical activity also raises FC-mediated AD risk, in line with studies showing exercise protects against AD through effects on amyloid *β*, inflammation, and blood flow (De la Rosa et al., 2020). Time spent outdoors, in both summer and winter, appears strongly protective, whereas TV viewing shows little effect. Longer sleep duration increases risk, supporting findings associating over nine hours of sleep with doubled AD risk (Westwood et al., 2017). Better self-rated health also lowers FC-mediated risk.

Among physiological factors, higher pulse rate and diabetes diagnosis significantly increase AD risk, consistent with links between poor glucose control and earlier dementia onset (Barbiellini Amidei et al., 2021; Imahori et al., 2022; Nguyen et al., 2020; Stanciu et al., 2020). The asymmetry in diabetes intervention effects likely reflects the low prevalence of diabetes in the dataset. Increased waist circumference raises AD risk, despite lower weight often being observed in diagnosed AD patients (Hsu et al., 2016; S. Singh et al., 1988). In cognitively normal subjects, however, higher weight and BMI correlate with AD-like FC patterns, especially in younger cohorts (e.g., HCP dataset), suggesting an age-dependent relationship (Gu et al., 2014). Lower red blood cell (RBC) count and hemoglobin levels are associated with greater AD risk, matching studies that identify reduced hemoglobin as a risk factor (W.-L. Chen et al., 2022; J. W. Kim et al., 2021; Wolters et al., 2019). Interestingly, both high and low blood pressure increase AD risk, revealing a U-shaped relationship. While midlife hypertension is a known AD risk factor (Sierra, 2020), blood pressure variability and hypotension are also implicated (Ebinger et al., 2023; Landin et al., 1993; Ma et al., 2024). To our knowledge, this U-shaped pattern is a novel finding. Lastly, hand grip strength has a minor effect but trends toward a protective role, consistent with evidence linking muscle strength to reduced dementia risk (Cui et al., 2021).

#### 4.2.2 Earlier-Life Factors Leveraging HCP Dataset

Figure 5 shows FC-mediated AD risk changes from interventions on genetic, modifiable, and physiological factors in the HCP dataset. While overall trends align with the UKB analysis, notable differences emerge, especially given that HCP subjects are over 30 years younger on average.

**Figure 5:**
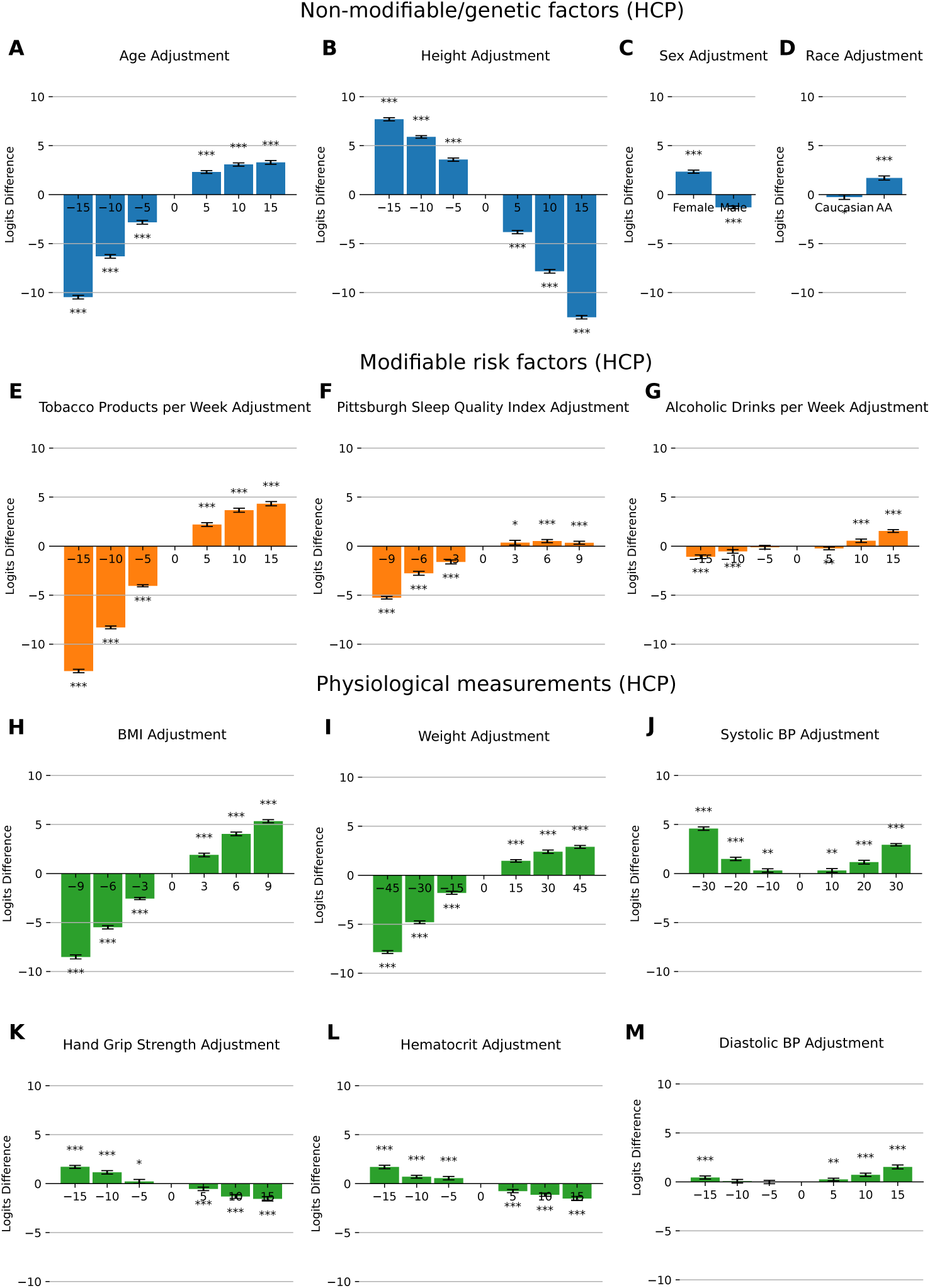
Changes in AD risk as quantified by FC mediator associated with non-modifiable/genetic factors, modifiable risk factors, and physiological measures in the HCP dataset. Fields are ordered by magnitude of change in AD risk. See caption of Figure 4 for additional details. Refer to Supplementary Materials Table 7 for summary statistics and units.

Age has a larger impact in HCP than UKB, likely because the same absolute age adjustment represents a greater proportional change for younger individuals. Decreasing age significantly lowers AD risk, while increasing it has a smaller effect. Height and sex show similar AD risk effects as in UKB, with shorter stature and female sex associated with higher risk—supporting genetic links between height-related pathways and neurodegeneration (Crews et al., 2010; Jørgensen et al., 2020; Larsson et al., 2017; Nelson et al., 2015). Race also influences AD risk: African American subjects show slightly higher FC-mediated risk than Caucasians, comparable in magnitude to the male–female difference, consistent with prior findings (Barnes & Bennett, 2014).

Among modifiable factors, tobacco use again strongly increases AD risk. The HCP dataset also includes sleep quality (PSQI), revealing that better sleep is protective, though poor sleep has a less pronounced negative effect. This aligns with prior studies linking sleep disruption to AD (Gaur et al., 2022). Alcohol consumption shows minimal impact on AD risk, echoing mixed findings in the literature (Seemiller et al., 2024). In both datasets, weight and BMI show a strong relationship with AD risk, supporting findings that obesity at any age may elevate AD susceptibility (Gu et al., 2014; Whitmer et al., 2005). Blood pressure again demonstrates a U-shaped effect, both high and low values increase AD risk, suggesting that deviation from normal ranges may be harmful even in healthy populations. Hand grip strength remains a minor but consistent protective factor, consistent with its use as a biomarker for cognitive decline (Shaughnessy et al., 2020). Finally, higher hematocrit levels (as with RBC in UKB) are associated with reduced AD risk, while anemia increases it.

### 4.3 Validation of the VAE Model in Recapitulating FC Changes

To check the validity of synthetic samples generated by the VAE model, we tested the difference between extremes of three phenotypes in the UKB and HCP datasets: age, sex, and BMI. The resulting mean FC changes between groups for old minus young (age), male minus female (sex), and high BMI vs low BMI groups are shown in Figure 6. The synthetic samples were created using the same demographic information as real samples, with the division between groups as in Table 1.

**Figure 6:**
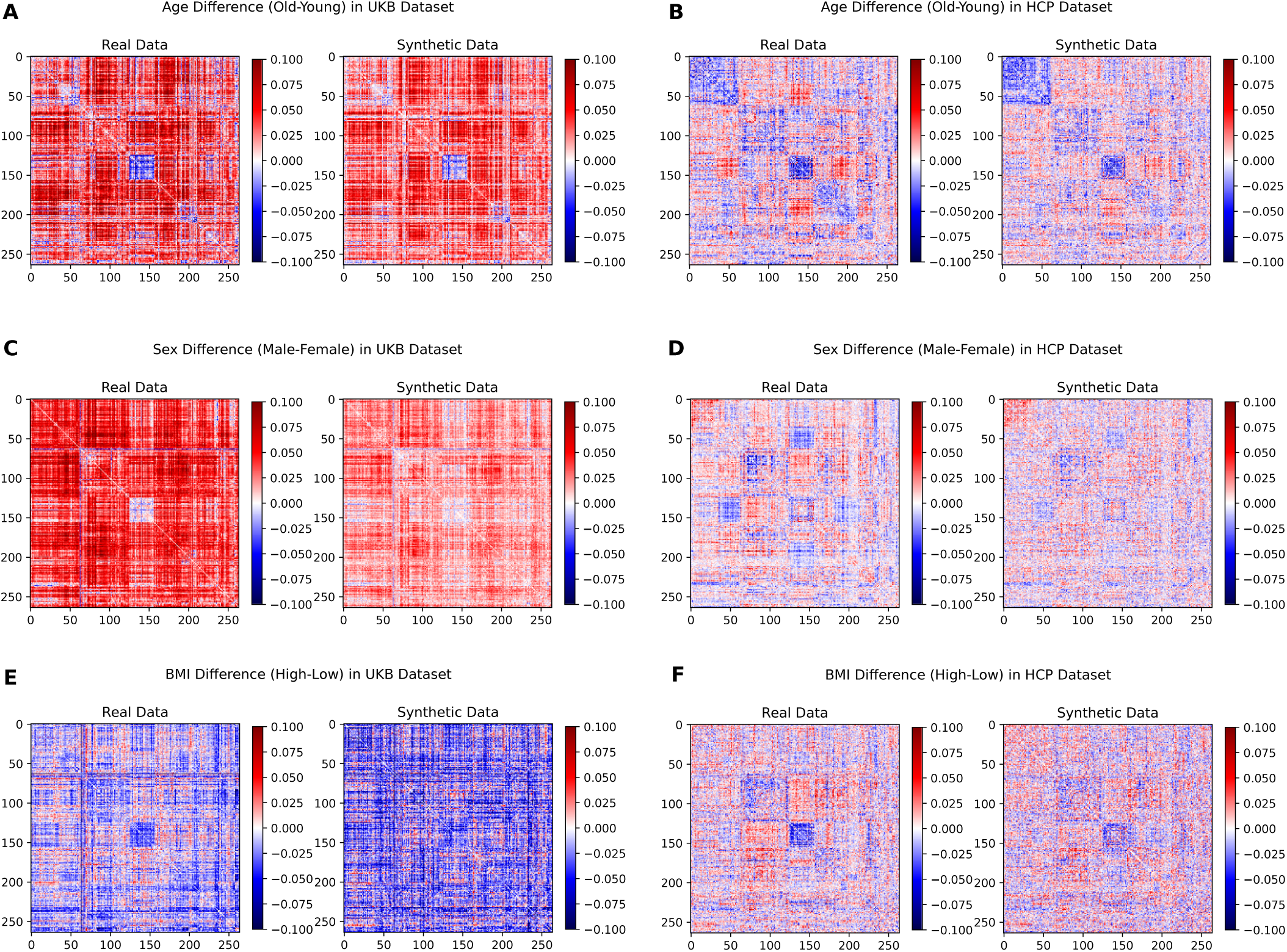
Changes in FC for real subjects groups (left of each pair) and synthetic subject groups (right of each pair) for differences in age (A,B), sex (C,D) and BMI (E,F) in the UKB and HCP datasets. We note that the synthetic, VAE-generated samples qualitatively reproduce group differences. It is also apparent that normative FC changes for the same demographic (e.g. age) are different for the UKB and HCP datasets.

**Table 1:**
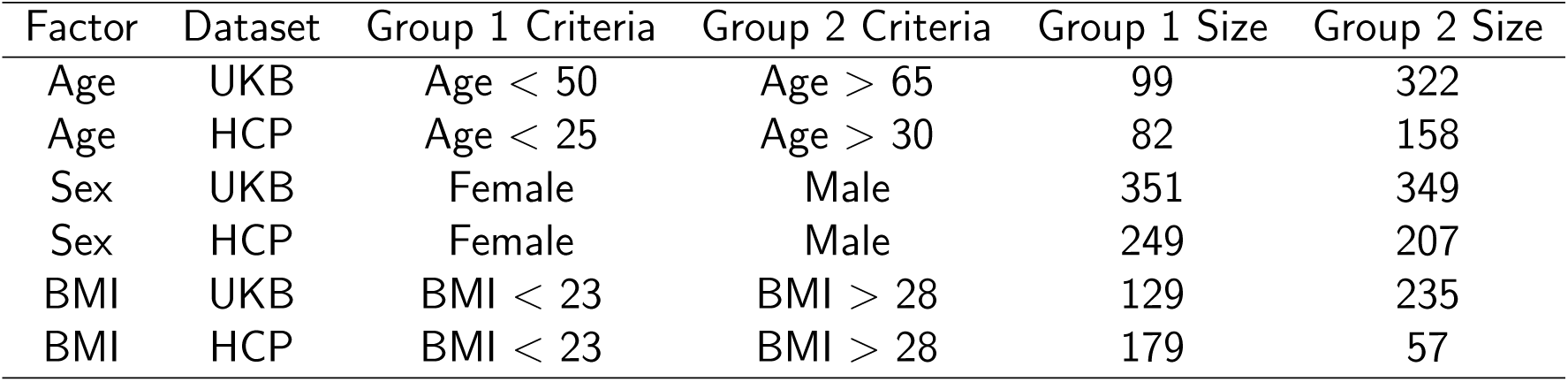
Group memberships for comparing real subject and synthetic sample group differences derived from the VAE model, as shown in Figure 6.

| Factor | Dataset | Group 1 Criteria | Group 2 Criteria | Group 1 Size | Group 2 Size |
| --- | --- | --- | --- | --- | --- |
| Age | UKB | Age < 50 | Age > 65 | 99 | 322 |
| Age | HCP | Age < 25 | Age > 30 | 82 | 158 |
| Sex | UKB | Female | Male | 351 | 349 |
| Sex | HCP | Female | Male | 249 | 207 |
| BMI | UKB | BMI < 23 | BMI > 28 | 129 | 235 |
| BMI | HCP | BMI < 23 | BMI > 28 | 179 | 57 |

The replication of group differences using synthetic samples leads us to believe the decoder accurately captures the effect of demographics or lifestyle factors on synthetically generated FC. Moreover, this highlights the main advantage of the VAE model over simply looking at group differences, i.e. the ability to intervene on one specific demographic/factor and keep all others the same. This has the effect of disentangling demographics that are otherwise correlated, e.g. age and height, sex and height, sex and weight, etc. The built-in disentangling effects of different demographics is one of the key benefits of using the VAE model that is not otherwise possible by using existing group differences in a naive way, due to the correlations between demographics/factors.

Finally, the similarity of distributions between synthetic FC data created via intervention by VAE and real FC data can be visualized directly by means of a low-dimensional manifold projection such as t-SNE (van der Maaten & Hinton, 2008). Figure 7 shows the t-SNE of synthetic data and real data in the UKB and HCP datasets. It can be seen that there is no discernible difference between the t-SNE projection of real versus synthetic FC data for the UKB or HCP datasets.

**Figure 7:**
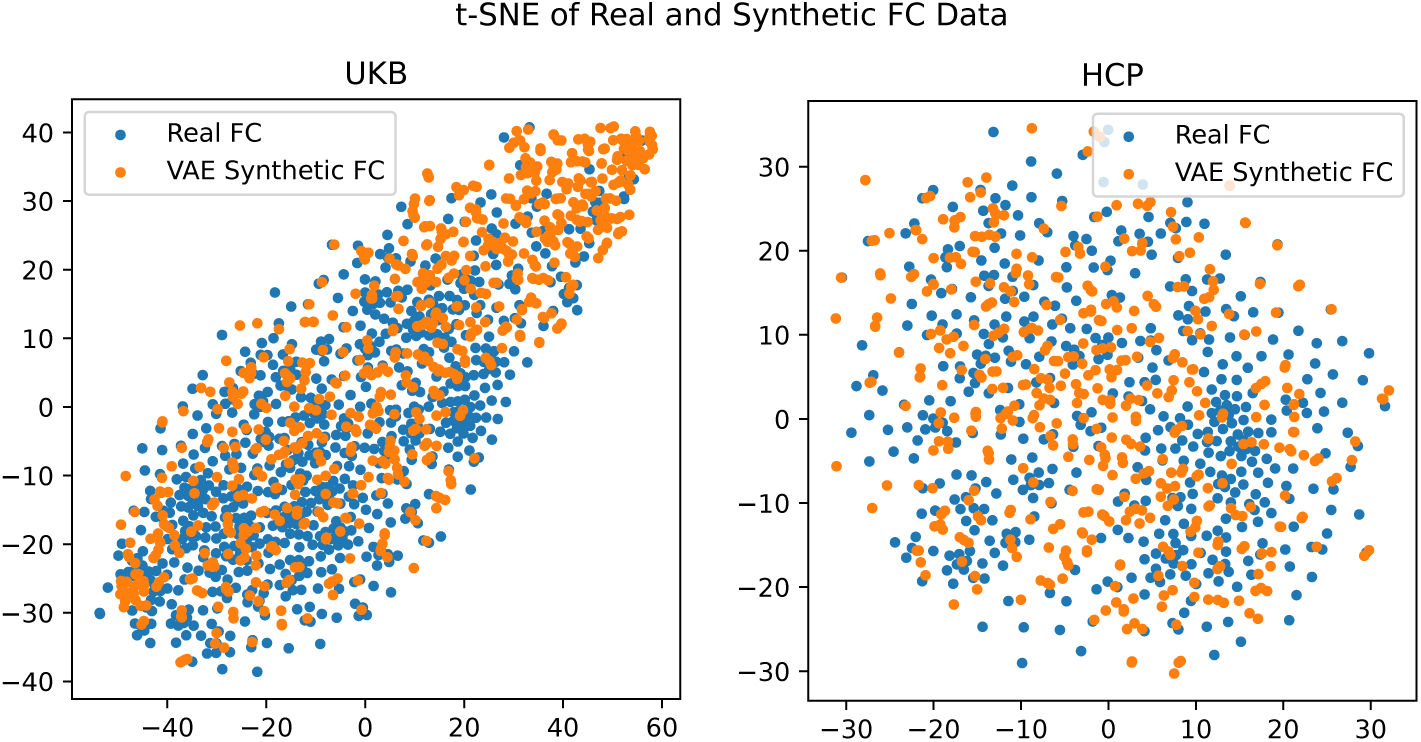
t-SNE projection of real and synthetic data from the UKB and HCP datasets. Shown are 700 subjects from the UKB and 456 subjects from the HCP datasets. Synthetic data had its latent space sampled from a multivariate normal distribution and was not a reconstruction of the original FC data. A perplexity of 10 was used to create the t-SNE.

### 4.4 Functional Connections Contributing to AD Risk Change

Understanding how interventions in the VAE model alter FC and contribute to AD risk is crucial. Figures 8 and 9 illustrate the impact of selected interventions (height, weight, waist circumference) on FC and AD risk per connection for the HCP and UKB datasets.

**Figure 8:**
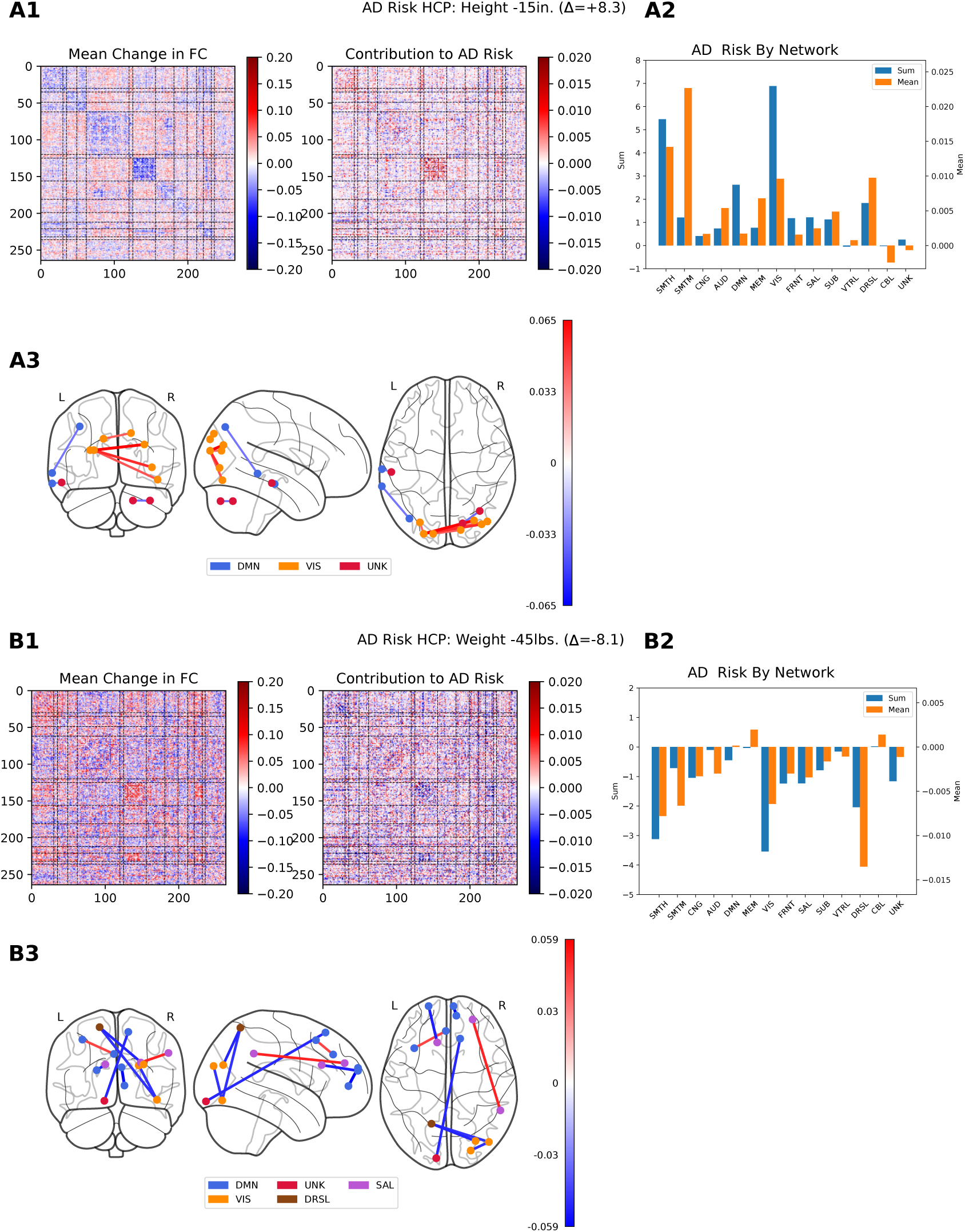
Selected changes in FC and concomitant change in AD risk profile for height (A) and weight (B) in the HCP dataset. Plots A1 and B1 show changes in FC for intervention to decrease height by 15 inches and decrease weight by 45 lbs. Plots A2 and B2 show average and total change in AD risk by brain functional network in the Power atlas. Plots A3 and B3 show most significant inter-ROI connections contributing to change in AD risk. We note that most significant connections for height are positive and in the VIS network, while connections for weight are negative and are long-range.

**Figure 9:**
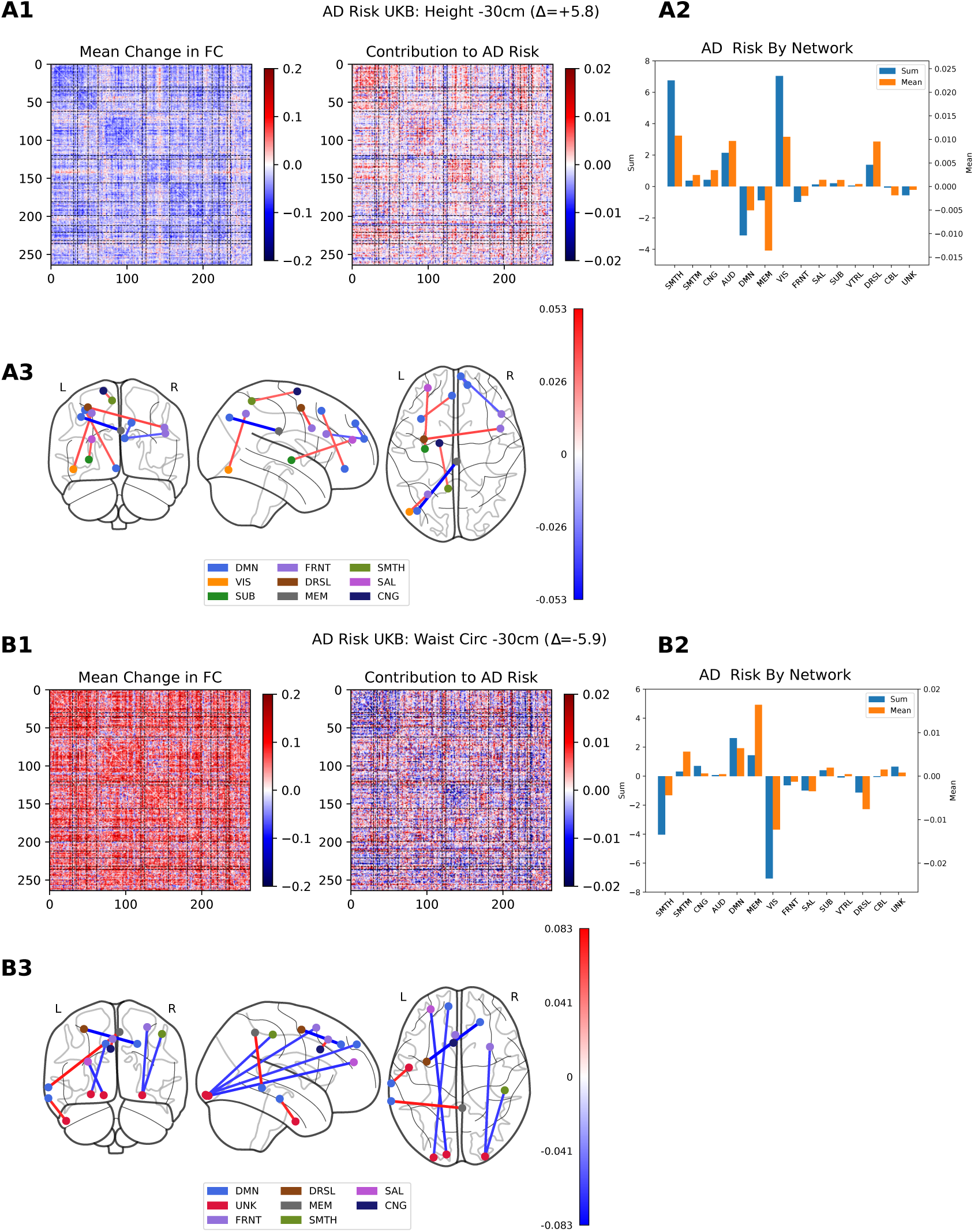
Selected changes in FC and concomitant change in AD risk profile for height (A) and waist circumference (B) intervention in the UKB dataset. Plots A1 and B1 show changes in FC for intervention to decrease height by 30 cm and decrease waist circumference by 30 cm. Plots A2 and B2 show average and total change in AD risk by brain functional network in the Power atlas. Plots A3 and B3 show most significant inter-ROI connections contributing to change in AD risk. Interventions on risk factors in the UKB cohort tend to increase or decrease FC across the whole connectome, as evident in this figure for an FC decrease in plot A1 and an FC increase in plot B1. This is in some contrast to HCP, where intervention tends to increase some inter-network connectivities while decreasing others.

In the HCP dataset (Figure 8), decreases in intra-network connectivity, particularly SMT-SMT and VIS-VIS, generally increase AD risk, while increases in these connections reduce it. SMT-VIS and other intra-network connections tend to co-vary, but DMN-VIS shows an opposing pattern. Despite varied FC changes, each network’s contribution to AD risk is mostly unidirectional, with the exception of the CBL network (Figures 8 A2 and B2). The strongest AD risk effects come from VIS, STMH, and to a lesser extent, DMN. Only the MEM, CBL, and UNK networks deviate from the overall risk trend, showing disagreement under certain interventions.

In contrast, the UKB dataset (Figure 9) shows more uniform FC changes across the connectome, either largely increasing or decreasing with intervention. This is especially evident for height and waist circumference. However, the corresponding AD risk varies by network. VIS and SMT again dominate in effect size, while DMN and MEM contribute smaller but oppositely directed effects. Unlike the HCP dataset, the UKB shows simpler global FC changes, but more diverse network-level AD risk responses.

## 5 Discussion

This project presents a novel framework for exploring how interventions on various phenotypic factors influence brain FC and, in turn, the risk of AD. By leveraging a phenotype-conditioned VAE, the method enables counterfactual manipulation of both modifiable and non-modifiable traits to assess their impact on FC patterns associated with AD. A model of FC changes characteristic of AD was first derived using the ADNI dataset. This was followed by the development of a VAE that conditions FC on individual covariates, allowing simulations of how altering specific traits would affect FC and AD risk. Key findings revealed that factors such as height, weight/BMI, tobacco use, and age (notably in the HCP dataset) significantly influenced FC-mediated AD risk. In the UKB dataset, higher polygenic risk scores were associated with increased risk, and a surprising U-shaped relationship was observed between blood pressure and AD risk across both the UKB and HCP datasets. Beyond AD, the approach provides a flexible tool for investigating the impact of phenotypic interventions on FC in other neurological disorders, such as schizophrenia or ADHD, by adapting the underlying risk model.

While our current study focuses on resting-state fMRI, the underlying framework is adaptable to other imaging modalities. Structural MRI, for example, has been extensively used in AD diagnosis, particularly in identifying atrophy patterns in later disease stages. Applying our model to sMRI or combining it with fMRI could enable multimodal inference across disease stages. Future work may also explore task-based fMRI, which has shown improved sensitivity in predicting cognitive traits and clinical outcomes compared to resting-state data in some studies (Zhao et al., 2023). Evaluating both modalities within the same cohort could clarify their respective contributions to early AD risk modeling and assess the consistency of inferred risk trajectories.

Another natural extension of this work is to incorporate dynamic FC, which captures time-varying changes in brain connectivity and may better reflect the brain’s adaptive processes than static FC. Dynamic FC has shown promise in improving classification of neuropsychiatric conditions such as schizophrenia (Rashid et al., 2016), and may enhance the sensitivity of AD risk modeling to transient or subtle network disruptions. Investigating whether dynamic FC yields consistent or complementary findings compared to static FC would help deepen mechanistic insights into how early-life factors shape long-term brain function and disease susceptibility. These future directions will further validate and extend the versatility of our approach across modalities and temporal dimensions.

Finally, although the present study performs a rigorous integration of early-life behaviors and risk factors and late-life AD risk, it would be valuable to obtain a large ground truth dataset, similar to the UKB but enrolling subjects at a younger age and following them until prodromal AD symptoms appear. Given the necessarily long time and resources required for this study, it is probably only possible with the cooperation of a national healthcare system. Even if unlikely in the near future, such a study would be the gold standard for the investigation of early-life risk factors and progression of AD in later life.

## Supporting information

Supplementary Analysis

## Data and Code Availability

ADNI, HCP, and UKB data are available from the relevant organizations to authorized researchers. The authors do not have permission to redistribute data. Model source code is made available at https://github.com/aorliche/InterventionVAE.

## Author Contributions

**Anton Orlichenko**: conceptualization, data curation, formal analysis, investigation, methodology, software, visualization, writing - original draft. **Shengxian Ding**: data curation, methodology, writing - review and editing. **Emily Johns**: investigation, software, methodology, writing - review and editing. **Zhiling Gu**: methodology, visualization, writing - review and editing. **Xinyuan Tian**: data curation, software. **Xiaoxuan Li**: investigation, writing - review and editing. **Yize Zhao**: conceptualization, funding acquisition, supervision, writing - review and editing.

## Funding

This work was partially supported by the National Institutes of Health under awards RF1AG081413, R01EB034720 and R01AG068191.

## Declaration of Competing Interests

The authors report no competing interests.

## Supplementary Material

### Correlation of FC and Clinical Fields in the UKB Dataset

Prior to VAE-based intervention analysis on modifiable and physiological factors, a preliminary study was undertaken to find phenotypic fields that were significantly correlated with FC in the UKB dataset. This was accomplished in two steps. First, treating each functional connection as an independent scalar measurement for each subject, the correlation between each functional connection and phenotypic field was calculated. Next, using the resulting correlation map with FC, the dot product of each subjects vectorized FC and correlation map was produced, with a scalar value for each subject.

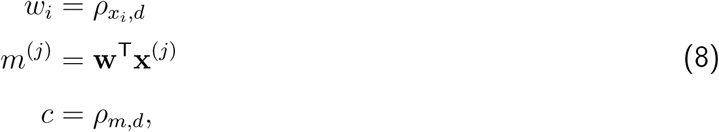

Here *x_i_* is the functional connection *i*, *d* is the phenotype of interest, 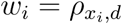 is the correlation between the phenotype of interest and a single functional connection among all subjects, *m*^(^*^j^*^)^ is the scalar dot product between the correlation map **w** and the vectorized FC **x**^(^*^j^*^)^ for subject *j*, and *c* is the final correlation. The correlation between this scalar dot product and phenotypic value was then calculated and this correlation is reported in Tables 2, 3, 4, and 5.

**Table 2:**
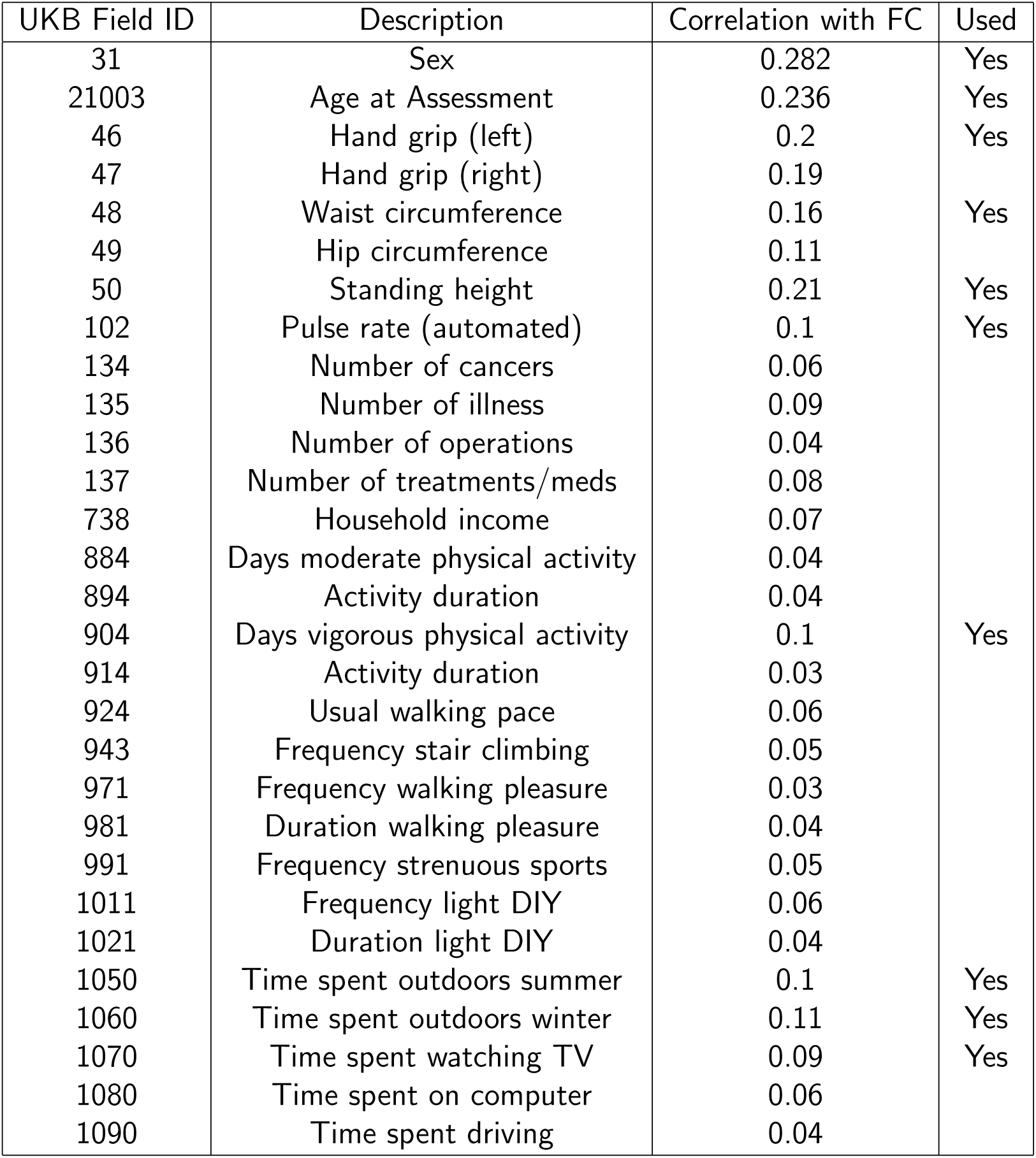
Correlations between UKB fields and resting state FC based on the Power atlas.

**Table 3:**
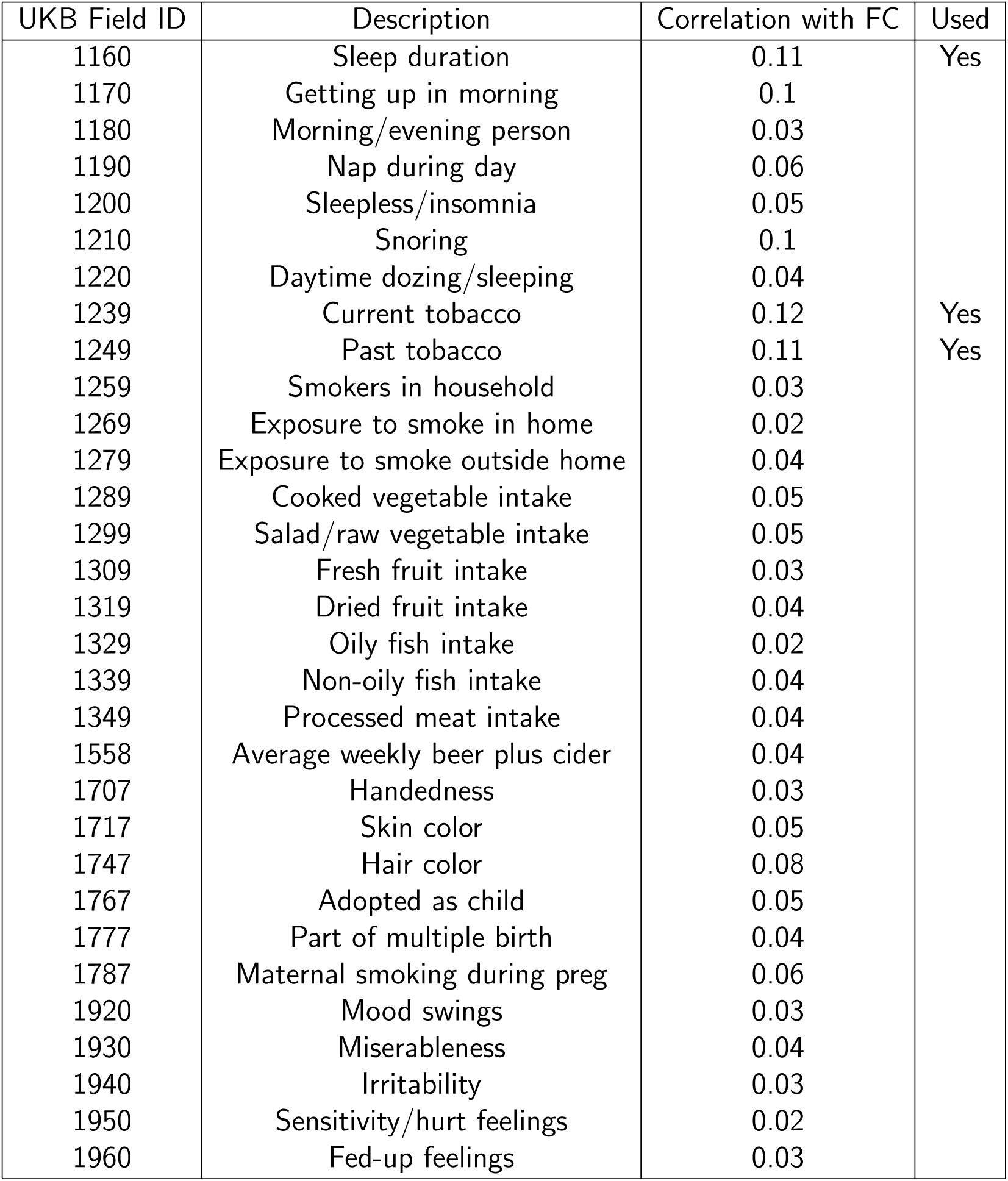
Correlation between UKB fields and resting state FC based on the Power atlas, continued.

**Table 4:**
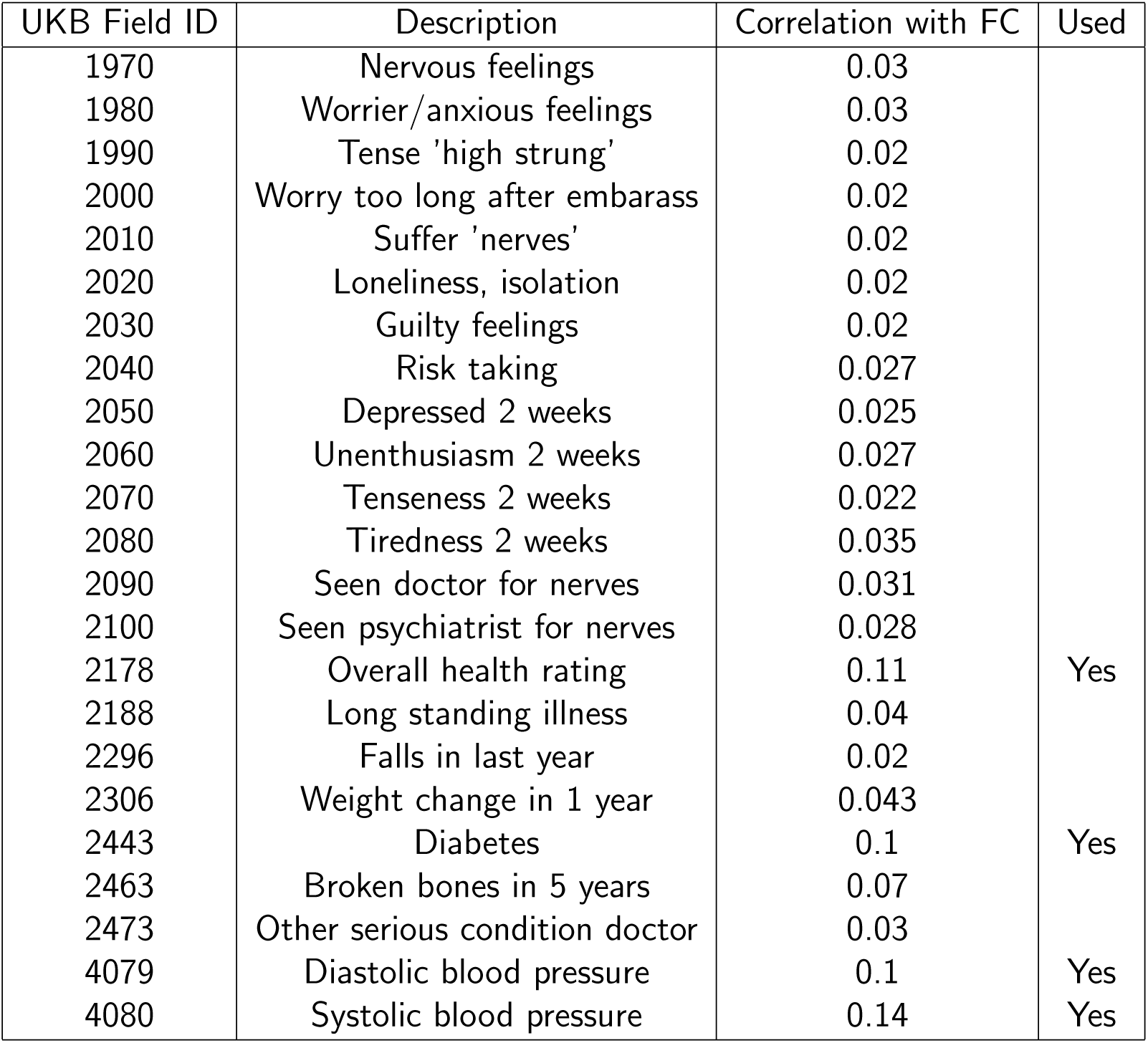
Correlation between UKB fields and resting state FC based on the Power atlas, continued.

**Table 5:**
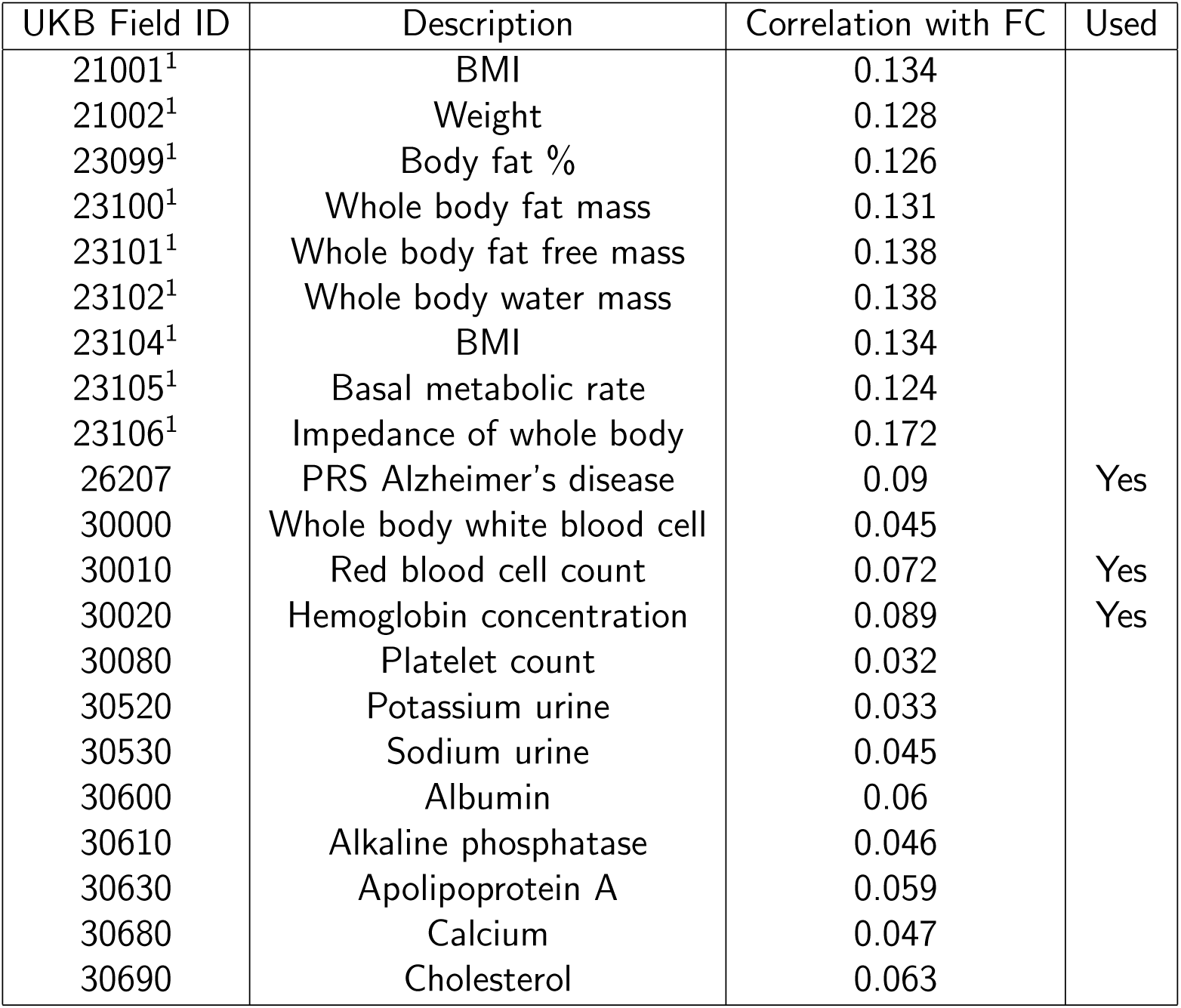
Correlation between UKB fields and resting state FC based on the Power atlas, continued. ^1^BMI, weight, and related fields were omitted as being redundant with waist circumference.

### UKB and HCP Intervention Fields

**Table 6:**
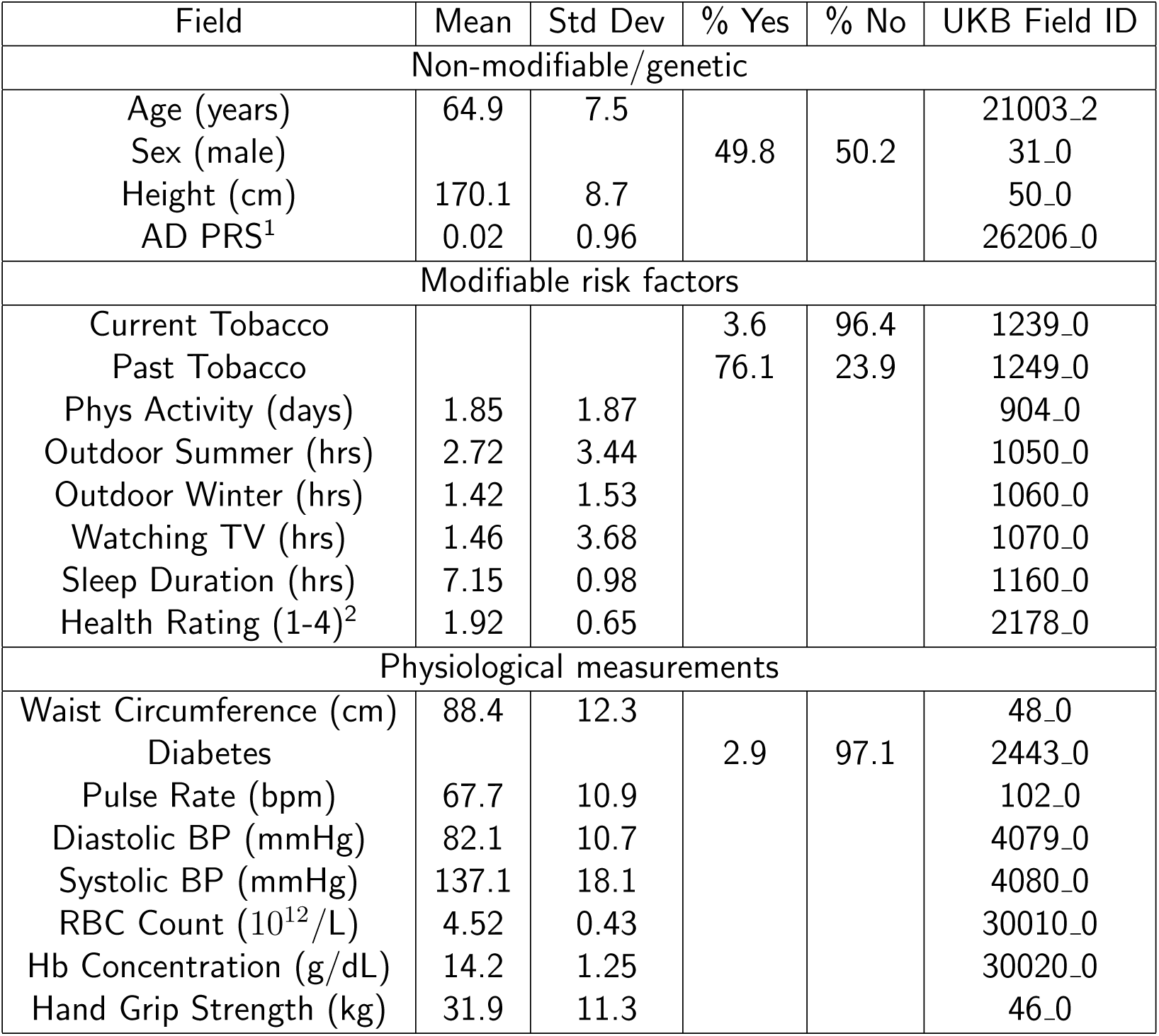
Non-modifiable/genetic factors, modifiable lifestyle factors, and physiologic measurements from the UKB dataset included in the VAE model. ^1^Polygenic risk score. ^2^Lower health rating is better.

**Table 7:**
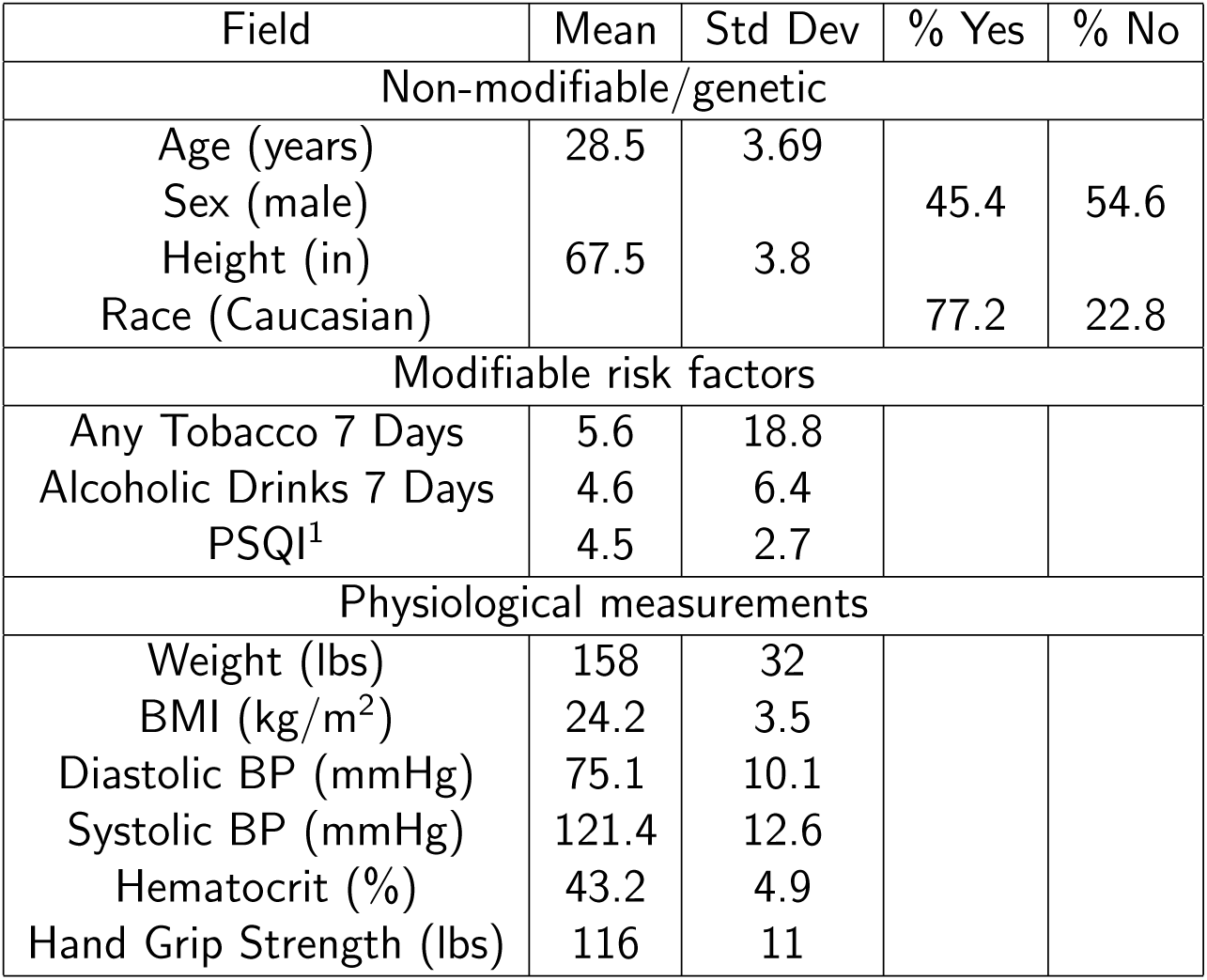
Non-modifiable/genetic factors, modifiable lifestyle factors, and physiological measurements from the HCP dataset included in the VAE model. ^1^Pittsburgh Sleep Quality Index; lower scores better.

### Top Positively and Negatively Weighted Connections for AD Risk

**Table 8:**
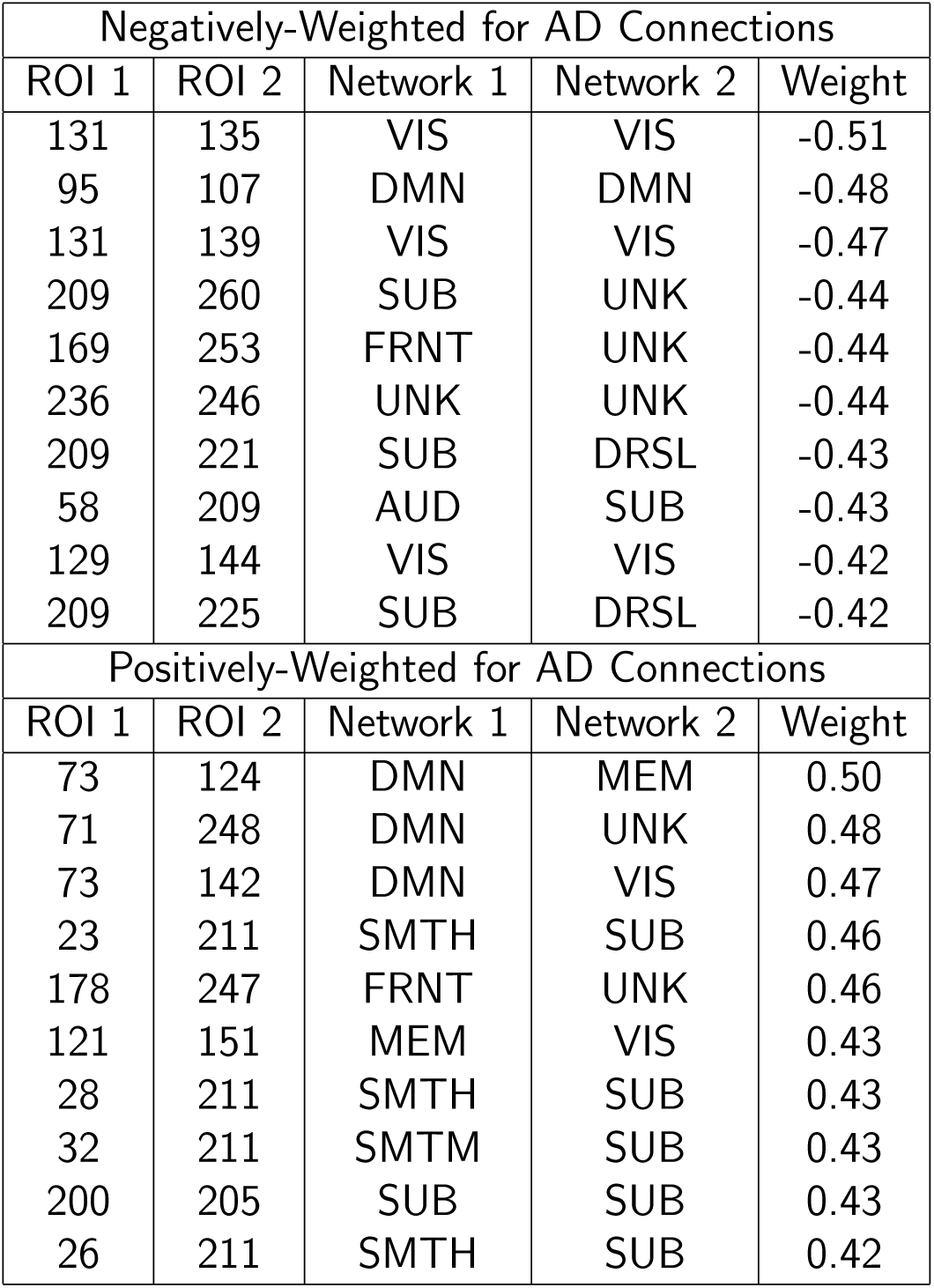
List of top ten connection weights positively and negatively correlated with AD in the aggregated ADNI AD risk model. ROIs are numbered starting from zero in the Power atlas. Note the large number of VIS network connections negatively correlated with AD risk. In contrast, there are many SUB and DMN connections with positive correlation with AD risk.

### Functional Networks in the Power Atlas

**Table 9:**
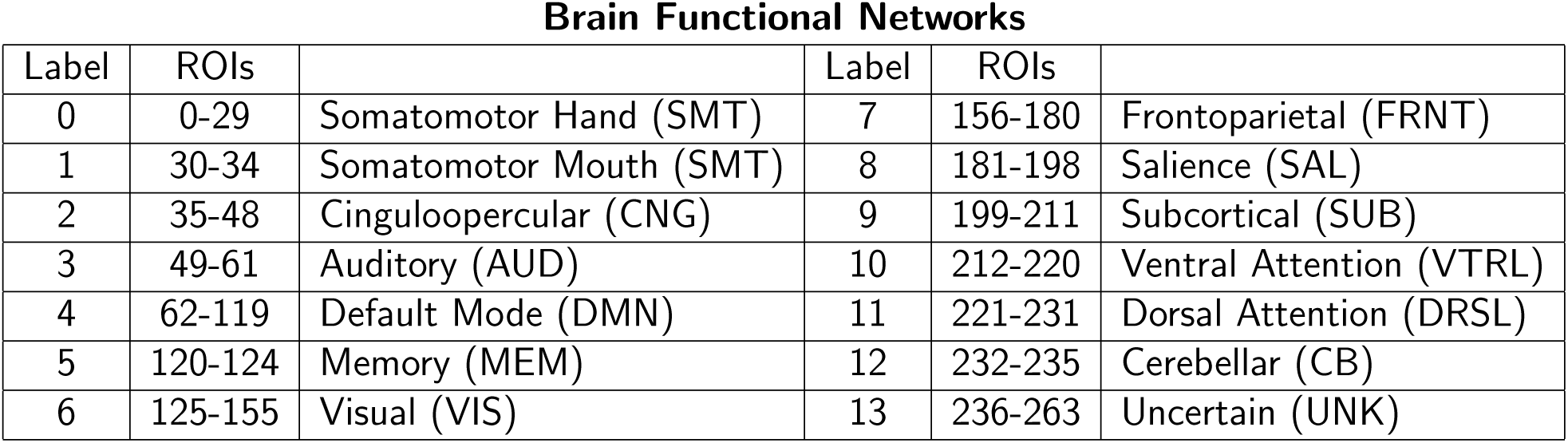
Regions, abbreviations, and functional networks in the Power atlas.

### Simulation Experiments

The ability of our counterfactual VAE model to accurately uncover the relationships between latent factors, an intermediate phenotype resembling FC, and outcome risk was tested in simulation experiments with a known ground truth. The simulation consisted of 6 latent factors, labeled *a* through *f*, which affected both the intermediate phenotype (**x**, **y**) and outcome risk *y*. In order to match our real FC-based analysis, two synthetic datasets, 𝒟*_X_* and 𝒟*_Y_*, with 𝒟*_X_* consisting of 1000 samples and 𝒟*_Y_* consisting of 100 samples, were created. Outcome risk *y* was known only for the 𝒟*_Y_* samples, however both **x** and **y** were the same functions of the latent factors. The synthetic data was instantiated as follows:

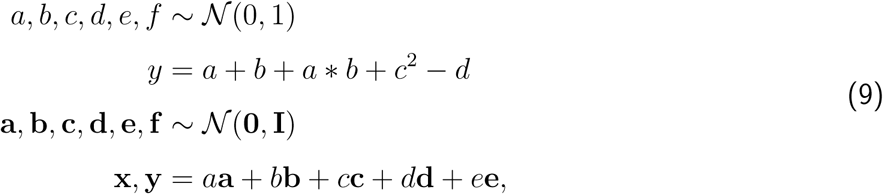

where **x** and **y** have 1000 features (**x**, **y** ∈ ℝ^1000^) and are functions of *a, b, c, d, e*, while the outcome *y* (a scalar) is a non-linear function of *a, b, c, d*. FC principal components **a**, **b**, **c**, **d**, **e**, **f** ∈ ℝ^1000^ are sampled once, and re-used for every synthetic subject, while a new sample of *a, b, c, d, e, f* is created for every subject. For the simulation experiments, we assume that *a, b, c, f* are observed latent factors while *d, e* are unobserved. Therefore both **x**, **y** and *y* are functions of both observed and unobserved latent factors.

Our goal in the simulation experiments was to determine the relationship between the observed latent factors *a, b, c, f* and outcome risk *y*. We performed this in two ways. First, as a baseline, we regressed **y** on observed features *a, b, c, f*, to determine the approximate values of *a, b, c, f* for each sample based on the relationship between *a, b, c, f* and **a**, **b**, **c**, **f** and **y**. Then, assuming outcome risk *y* was a second-order polynomial in *a, b, c, f*, we regressed the outcome risk *y* on the coefficients determined in the previous regression, thereby estimating the functional relationship between *y* and the latent factors *a, b, c, f*. Regression was performed using the Ridge model of scikit-learn package using three different values of the regularization parameter *α* Pedregosa et al., 2011. The initial regression coefficients corresponding to **a** through **f** were estimated on the larger dataset 𝒟*_X_*, then evaluated for the dataset 𝒟*_Y_* for which outcome risk *y* was available.

After determining the coefficients using the baseline two-stage regression, we then attempted to determine the coefficients of outcome risk based on latent factors using the intervention VAE model. We began by fitting an MLP on 𝒟*_Y_* to predict *y* from **y**. The MLP was a simple two-layer function with a size 30 hidden layer. The MLP was trained for 2000 epochs using the Adam optimizer with a learning rate of 10^−4^ and L2 regularization of 10^−5^. Then, we trained the intervention VAE on the dataset 𝒟*_X_* conditioned upon observed latent factors *a, b, c, f*. Once the VAE model was trained, we intervened on each of the latent factors in turn and determine the change in counterfactual outcome risk *ŷ* compared to *y* before intervention. We then fit coefficients to each of the latent factors, assuming the true function is a second-degree polynomial in the latent factors. The mean squared error of the coefficients determined using VAE was then compared to the coefficients found using the two-stage regression approach outlined above. The synthetic data was simulated ten times and coefficients were estimated for each simulation.

#### Simulation Results

The results of our simulation experiments are shown in Figure 10. We find that due to the ability of the VAE to create a highly reliable model on the larger synthetic dataset 𝒟*_X_*, the ability to estimate coefficients for *y* from 𝒟*_Y_* was greatly enhanced. The VAE was able to correctly estimate those coefficients (*a, b, a* ∗ *b, c*^2^) that were non-zero and set to zero those coefficients which were observed but not important for outcome risk (*f*). On the other hand, the two-stage regression we were comparing against was highly dependent on the regularization hyperparameter used for the regression. Using a large regularization hyperparameter *α* = 100, the two-stage regression was able to set *f* to be correctly equal to zero, but the other coefficients were found to be too small, around or less than 0.5 instead of the ground truth of one. Decreasing the regularization parameter allowed the coefficients of *a, b, a* ∗ *b, c*^2^ to reach their correct values, but with the downside that *f* was no longer exactly zero but had a distribution of values around zero. In any case, we found the variance of the coefficients estimated using two-stage regression to be much larger than for the interventional VAE model. Mean squared error for the coefficients for the VAE model and each two-stage regression are given in Table 10.

**Figure 10:**
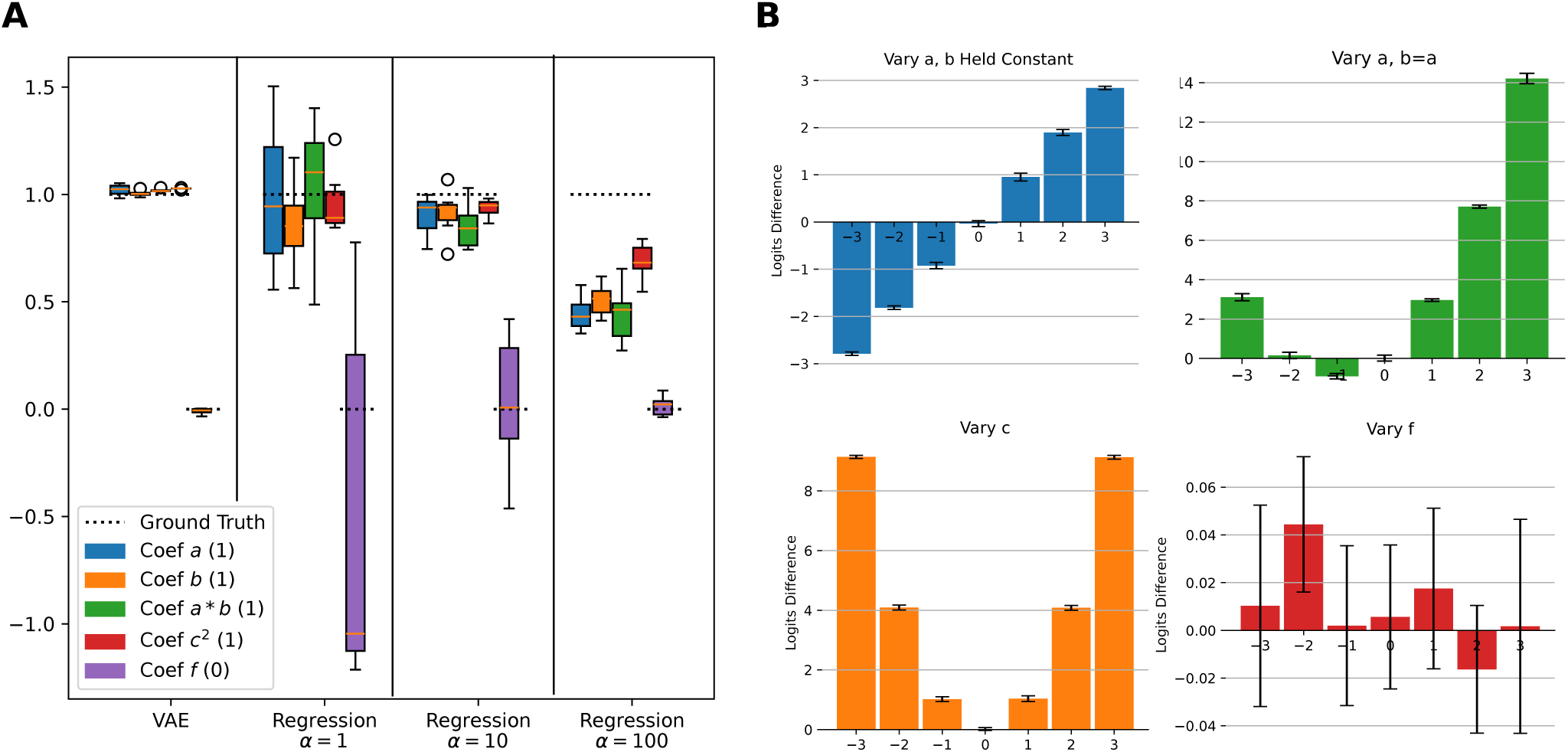
Results of the simulation experiments comparing use of the intervention VAE to two-stage regression in order to find the coefficients of a latent factor model of outcome risk. Part (A) shows estimates of coefficients for *a, b, a* ∗ *b, c*^2^, *f* using either VAE or two-stage regression at three different settings for the regularization hyperparameter *α*. It can be seen that the VAE is able to estimate coefficient values almost exactly while the two-stage regression has more variance and is highly dependent on the value of the regularization hyperparameter. (B) shows the method of estimating coefficients in the VAE model by intervening on each coefficient value and observing the resulting change in outcome risk.

**Table 10:**
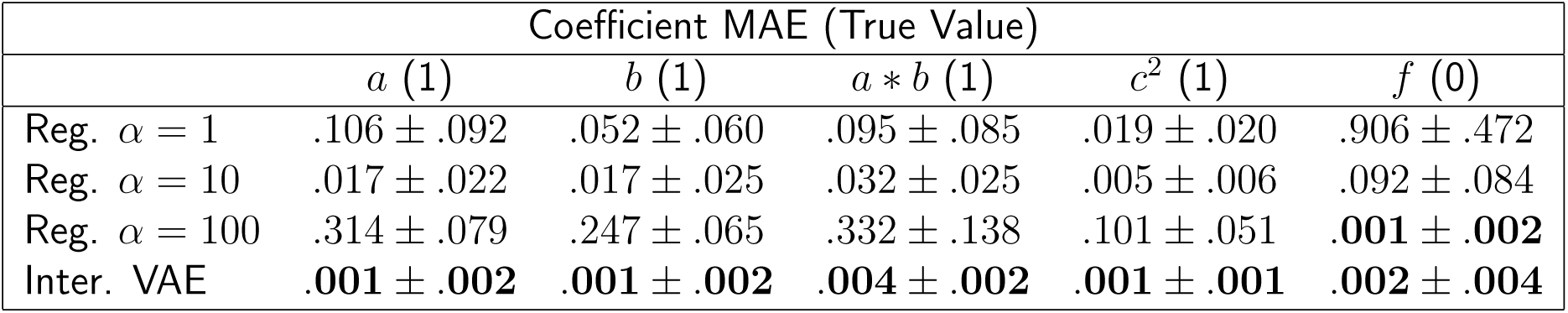
Results of simulation experiments. Mean squared error for coefficients of latent factors leading to outcome risk determined using the intervention VAE or two-stage regression model. Three regression models are shown, using three different settings of the L2 regularization hyperparameter *α*.

|  | Coefficient MAE (True Value) |  |  |  |  |
| --- | --- | --- | --- | --- | --- |
| | $a$ (1) | $b$ (1) | $a * b$ (1) | $c^2$ (1) | $f$ (0) |
| Reg. $\alpha = 1$ | .106 ± .092 | .052 ± .060 | .095 ± .085 | .019 ± .020 | .906 ± .472 |
| Reg. $\alpha = 10$ | .017 ± .022 | .017 ± .025 | .032 ± .025 | .005 ± .006 | .092 ± .084 |
| Reg. $\alpha = 100$ | .314 ± .079 | .247 ± .065 | .332 ± .138 | .101 ± .051 | **.001 ± .002** |
| Inter. VAE | **.001 ± .002** | **.001 ± .002** | **.004 ± .002** | **.001 ± .001** | **.002 ± .004** |

## Notes

### Competing Interest Statement

The authors have declared no competing interest.

