## Supplementary Analysis for "Intervention on Modifiable Lifestyle and Physiological Factors via Variational Autoencoder Reveals Changes in Functional Connectivity-Mediated Risk for Alzheimer’s Disease"

### 1. Correlation of FC and Clinical Fields in the UKB Dataset

Prior to VAE-based intervention analysis on modifiable and physiological factors, a preliminary study was undertaken to find phenotypic fields that were significantly correlated with FC in the UKB dataset. This was accomplished in two steps. First, treating each functional connection as an independent scalar measurement for each subject, the correlation between each functional connection and phenotypic field was calculated. Next, using the resulting correlation map with FC, the dot product of each subjects vectorized FC and correlation map was produced, with a scalar value for each subject.

$$\begin{aligned} w_i &= \rho_{x_i,d} \\ m^{(j)} &= \mathbf{w}^{\mathrm{T}}\mathbf{x}^{(j)} \\ c &= \rho_{m,d}, \end{aligned} \tag{1}$$

Here $x_i$ is the functional connection $i$, $d$ is the phenotype of interest, $w_i = \rho_{x_i,d}$ is the correlation between the phenotype of interest and a single functional connection among all subjects, $m^{(j)}$ is the scalar dot product between the correlation map $\mathbf{w}$ and the vectorized FC $\mathbf{x}^{(j)}$ for subject $j$, and $c$ is the final correlation. The correlation between this scalar dot product and phenotypic value was then calculated and this correlation is reported in Tables 1, 2, 3, and 4.

| UKB Field ID | Description | Correlation with FC | Used |
|---|---|---|---|
| 31 | Sex | 0.282 | Yes |
| 21003 | Age at Assessment | 0.236 | Yes |
| 46 | Hand grip (left) | 0.2 | Yes |
| 47 | Hand grip (right) | 0.19 | |
| 48 | Waist circumference | 0.16 | Yes |
| 49 | Hip circumference | 0.11 | |
| 50 | Standing height | 0.21 | Yes |
| 102 | Pulse rate (automated) | 0.1 | Yes |
| 134 | Number of cancers | 0.06 | |
| 135 | Number of illness | 0.09 | |
| 136 | Number of operations | 0.04 | |
| 137 | Number of treatments/meds | 0.08 | |
| 738 | Household income | 0.07 | |
| 884 | Days moderate physical activity | 0.04 | |
| 894 | Activity duration | 0.04 | |
| 904 | Days vigorous physical activity | 0.1 | Yes |
| 914 | Activity duration | 0.03 | |
| 924 | Usual walking pace | 0.06 | |
| 943 | Frequency stair climbing | 0.05 | |
| 971 | Frequency walking pleasure | 0.03 | |
| 981 | Duration walking pleasure | 0.04 | |
| 991 | Frequency strenuous sports | 0.05 | |
| 1011 | Frequency light DIY | 0.06 | |
| 1021 | Duration light DIY | 0.04 | |
| 1050 | Time spent outdoors summer | 0.1 | Yes |
| 1060 | Time spent outdoors winter | 0.11 | Yes |
| 1070 | Time spent watching TV | 0.09 | Yes |
| 1080 | Time spent on computer | 0.06 | |
| 1090 | Time spent driving | 0.04 | |

Table 1: Correlations between UKB fields and resting state FC based on the Power atlas.

| UKB Field ID | Description | Correlation with FC | Used |
|---|---|---|---|
| 1160 | Sleep duration | 0.11 | Yes |
| 1170 | Getting up in morning | 0.1 | |
| 1180 | Morning/evening person | 0.03 | |
| 1190 | Nap during day | 0.06 | |
| 1200 | Sleepless/insomnia | 0.05 | |
| 1210 | Snoring | 0.1 | |
| 1220 | Daytime dozing/sleeping | 0.04 | |
| 1239 | Current tobacco | 0.12 | Yes |
| 1249 | Past tobacco | 0.11 | Yes |
| 1259 | Smokers in household | 0.03 | |
| 1269 | Exposure to smoke in home | 0.02 | |
| 1279 | Exposure to smoke outside home | 0.04 | |
| 1289 | Cooked vegetable intake | 0.05 | |
| 1299 | Salad/raw vegetable intake | 0.05 | |
| 1309 | Fresh fruit intake | 0.03 | |
| 1319 | Dried fruit intake | 0.04 | |
| 1329 | Oily fish intake | 0.02 | |
| 1339 | Non-oily fish intake | 0.04 | |
| 1349 | Processed meat intake | 0.04 | |
| 1558 | Average weekly beer plus cider | 0.04 | |
| 1707 | Handedness | 0.03 | |
| 1717 | Skin color | 0.05 | |
| 1747 | Hair color | 0.08 | |
| 1767 | Adopted as child | 0.05 | |
| 1777 | Part of multiple birth | 0.04 | |
| 1787 | Maternal smoking during preg | 0.06 | |
| 1920 | Mood swings | 0.03 | |
| 1930 | Miserableness | 0.04 | |
| 1940 | Irritability | 0.03 | |
| 1950 | Sensitivity/hurt feelings | 0.02 | |
| 1960 | Fed-up feelings | 0.03 | |

Table 2: Correlation between UKB fields and resting state FC based on the Power atlas, continued.

| UKB Field ID | Description | Correlation with FC | Used |
|---|---|---|---|
| 1970 | Nervous feelings | 0.03 | |
| 1980 | Worrier/anxious feelings | 0.03 | |
| 1990 | Tense 'high strung' | 0.02 | |
| 2000 | Worry too long after embarass | 0.02 | |
| 2010 | Suffer 'nerves' | 0.02 | |
| 2020 | Loneliness, isolation | 0.02 | |
| 2030 | Guilty feelings | 0.02 | |
| 2040 | Risk taking | 0.027 | |
| 2050 | Depressed 2 weeks | 0.025 | |
| 2060 | Unenthusiasm 2 weeks | 0.027 | |
| 2070 | Tenseness 2 weeks | 0.022 | |
| 2080 | Tiredness 2 weeks | 0.035 | |
| 2090 | Seen doctor for nerves | 0.031 | |
| 2100 | Seen psychiatrist for nerves | 0.028 | |
| 2178 | Overall health rating | 0.11 | Yes |
| 2188 | Long standing illness | 0.04 | |
| 2296 | Falls in last year | 0.02 | |
| 2306 | Weight change in 1 year | 0.043 | |
| 2443 | Diabetes | 0.1 | Yes |
| 2463 | Broken bones in 5 years | 0.07 | |
| 2473 | Other serious condition doctor | 0.03 | |
| 4079 | Diastolic blood pressure | 0.1 | Yes |
| 4080 | Systolic blood pressure | 0.14 | Yes |

Table 3: Correlation between UKB fields and resting state FC based on the Power atlas, continued.

| UKB Field ID | Description | Correlation with FC | Used |
|---|---|---|---|
| 21001[1] | BMI | 0.134 | |
| 21002[1] | Weight | 0.128 | |
| 23099[1] | Body fat % | 0.126 | |
| 23100[1] | Whole body fat mass | 0.131 | |
| 23101[1] | Whole body fat free mass | 0.138 | |
| 23102[1] | Whole body water mass | 0.138 | |
| 23104[1] | BMI | 0.134 | |
| 23105[1] | Basal metabolic rate | 0.124 | |
| 23106[1] | Impedance of whole body | 0.172 | |
| 26207 | PRS Alzheimer's disease | 0.09 | Yes |
| 30000 | Whole body white blood cell | 0.045 | |
| 30010 | Red blood cell count | 0.072 | Yes |
| 30020 | Hemoglobin concentration | 0.089 | Yes |
| 30080 | Platelet count | 0.032 | |
| 30520 | Potassium urine | 0.033 | |
| 30530 | Sodium urine | 0.045 | |
| 30600 | Albumin | 0.06 | |
| 30610 | Alkaline phosphatase | 0.046 | |
| 30630 | Apolipoprotein A | 0.059 | |
| 30680 | Calcium | 0.047 | |
| 30690 | Cholesterol | 0.063 | |

Table 4: Correlation between UKB fields and resting state FC based on the Power atlas, continued. [1]BMI, weight, and related fields were omitted as being redundant with waist circumference.

### 2. UKB and HCP Intervention Fields

| Field | Mean | Std Dev | % Yes | % No | UKB Field ID |
|---|---|---|---|---|---|
| Non-modifiable/genetic | | | | | |
| Age (years) | 64.9 | 7.5 | | | 21003_2 |
| Sex (male) | | | 49.8 | 50.2 | 31_0 |
| Height (cm) | 170.1 | 8.7 | | | 50_0 |
| AD PRS[1] | 0.02 | 0.96 | | | 26206_0 |
| Modifiable risk factors | | | | | |
| Current Tobacco | | | 3.6 | 96.4 | 1239_0 |
| Past Tobacco | | | 76.1 | 23.9 | 1249_0 |
| Phys Activity (days) | 1.85 | 1.87 | | | 904_0 |
| Outdoor Summer (hrs) | 2.72 | 3.44 | | | 1050_0 |
| Outdoor Winter (hrs) | 1.42 | 1.53 | | | 1060_0 |
| Watching TV (hrs) | 1.46 | 3.68 | | | 1070_0 |
| Sleep Duration (hrs) | 7.15 | 0.98 | | | 1160_0 |
| Health Rating (1-4)[2] | 1.92 | 0.65 | | | 2178_0 |
| Physiological measurements | | | | | |
| Waist Circumference (cm) | 88.4 | 12.3 | | | 48_0 |
| Diabetes | | | 2.9 | 97.1 | 2443_0 |
| Pulse Rate (bpm) | 67.7 | 10.9 | | | 102_0 |
| Diastolic BP (mmHg) | 82.1 | 10.7 | | | 4079_0 |
| Systolic BP (mmHg) | 137.1 | 18.1 | | | 4080_0 |
| RBC Count ($10^{12}$/L) | 4.52 | 0.43 | | | 30010_0 |
| Hb Concentration (g/dL) | 14.2 | 1.25 | | | 30020_0 |
| Hand Grip Strength (kg) | 31.9 | 11.3 | | | 46_0 |

Table 5: Non-modifiable/genetic factors, modifiable lifestyle factors, and physiologic measurements from the UKB dataset included in the VAE model. [1]Polygenic risk score. [2]Lower health rating is better.

| Field | Mean | Std Dev | % Yes | % No |
|---|---|---|---|---|
| Non-modifiable/genetic | | | | |
| Age (years) | 28.5 | 3.69 | | |
| Sex (male) | | | 45.4 | 54.6 |
| Height (in) | 67.5 | 3.8 | | |
| Race (Caucasian) | | | 77.2 | 22.8 |
| Modifiable risk factors | | | | |
| Any Tobacco 7 Days | 5.6 | 18.8 | | |
| Alcoholic Drinks 7 Days | 4.6 | 6.4 | | |
| PSQI[1] | 4.5 | 2.7 | | |
| Physiological measurements | | | | |
| Weight (lbs) | 158 | 32 | | |
| BMI ($kg/m^2$) | 24.2 | 3.5 | | |
| Diastolic BP (mmHg) | 75.1 | 10.1 | | |
| Systolic BP (mmHg) | 121.4 | 12.6 | | |
| Hematocrit (%) | 43.2 | 4.9 | | |
| Hand Grip Strength (lbs) | 116 | 11 | | |

Table 6: Non-modifiable/genetic factors, modifiable lifestyle factors, and physiological measurements from the HCP dataset included in the VAE model. [1]Pittsburgh Sleep Quality Index; lower scores better.

### 3. Top Positively and Negatively Weighted Connections for AD Risk

| Negatively-Weighted for AD Connections | | | | |
|---|---|---|---|---|
| ROI 1 | ROI 2 | Network 1 | Network 2 | Weight |
| 131 | 135 | VIS | VIS | -0.51 |
| 95 | 107 | DMN | DMN | -0.48 |
| 131 | 139 | VIS | VIS | -0.47 |
| 209 | 260 | SUB | UNK | -0.44 |
| 169 | 253 | FRNT | UNK | -0.44 |
| 236 | 246 | UNK | UNK | -0.44 |
| 209 | 221 | SUB | DRSL | -0.43 |
| 58 | 209 | AUD | SUB | -0.43 |
| 129 | 144 | VIS | VIS | -0.42 |
| 209 | 225 | SUB | DRSL | -0.42 |
| Positively-Weighted for AD Connections | | | | |
| ROI 1 | ROI 2 | Network 1 | Network 2 | Weight |
| 73 | 124 | DMN | MEM | 0.50 |
| 71 | 248 | DMN | UNK | 0.48 |
| 73 | 142 | DMN | VIS | 0.47 |
| 23 | 211 | SMTH | SUB | 0.46 |
| 178 | 247 | FRNT | UNK | 0.46 |
| 121 | 151 | MEM | VIS | 0.43 |
| 28 | 211 | SMTH | SUB | 0.43 |
| 32 | 211 | SMTM | SUB | 0.43 |
| 200 | 205 | SUB | SUB | 0.43 |
| 26 | 211 | SMTH | SUB | 0.42 |

Table 7: List of top ten connection weights positively and negatively correlated with AD in the aggregated ADNI AD risk model. ROIs are numbered starting from zero in the Power atlas. Note the large number of VIS network connections negatively correlated with AD risk. In contrast, there are many SUB and DMN connections with positive correlation with AD risk.

### 4. Functional Networks in the Power Atlas

**Brain Functional Networks**

| Label | ROIs | | Label | ROIs | |
|---|---|---|---|---|---|
| 0 | 0-29 | Somatomotor Hand (SMT) | 7 | 156-180 | Frontoparietal (FRNT) |
| 1 | 30-34 | Somatomotor Mouth (SMT) | 8 | 181-198 | Salience (SAL) |
| 2 | 35-48 | Cinguloopercular (CNG) | 9 | 199-211 | Subcortical (SUB) |
| 3 | 49-61 | Auditory (AUD) | 10 | 212-220 | Ventral Attention (VTRL) |
| 4 | 62-119 | Default Mode (DMN) | 11 | 221-231 | Dorsal Attention (DRSL) |
| 5 | 120-124 | Memory (MEM) | 12 | 232-235 | Cerebellar (CB) |
| 6 | 125-155 | Visual (VIS) | 13 | 236-263 | Uncertain (UNK) |

Table 8: Regions, abbreviations, and functional networks in the Power atlas.

### 5. Simulation Experiments

The ability of our counterfactual VAE model to accurately uncover the relationships between latent factors, an intermediate phenotype resembling FC, and outcome risk was tested in simulation experiments with a known ground truth. The simulation consisted of 6 latent factors, labeled $a$ through $f$, which affected both the intermediate phenotype ($\mathbf{x}$, $\mathbf{y}$) and outcome risk $y$. In order to match our real FC-based analysis, two synthetic datasets, $\mathcal{D}_X$ and $\mathcal{D}_Y$, with $\mathcal{D}_X$ consisting of 1000 samples and $\mathcal{D}_Y$ consisting of 100 samples, were created. Outcome risk $y$ was known only for the $\mathcal{D}_Y$ samples, however both $\mathbf{x}$ and $\mathbf{y}$ were the same functions of the latent factors. The synthetic data was instantiated as follows:

$$
\begin{aligned}
a, b, c, d, e, f &\sim \mathcal{N}(0, 1) \\
y &= a + b + a * b + c^2 - d \\
\mathbf{a}, \mathbf{b}, \mathbf{c}, \mathbf{d}, \mathbf{e}, \mathbf{f} &\sim \mathcal{N}(\mathbf{0}, \mathbf{I}) \\
\mathbf{x}, \mathbf{y} &= a\mathbf{a} + b\mathbf{b} + c\mathbf{c} + d\mathbf{d} + e\mathbf{e},
\end{aligned}
\tag{2}
$$

where $\mathbf{x}$ and $\mathbf{y}$ have 1000 features ($\mathbf{x}, \mathbf{y} \in \mathbb{R}^{1000}$) and are functions of $a, b, c, d, e$, while the outcome $y$ (a scalar) is a non-linear function of $a, b, c, d$. FC principal components $\mathbf{a}, \mathbf{b}, \mathbf{c}, \mathbf{d}, \mathbf{e}, \mathbf{f} \in \mathbb{R}^{1000}$ are sampled once, and re-used for every synthetic subject, while a new sample of $a, b, c, d, e, f$ is created for every subject. For the simulation experiments, we assume that $a, b, c, f$ are observed latent factors while $d, e$ are unobserved. Therefore both $\mathbf{x}, \mathbf{y}$ and $y$ are functions of both observed and unobserved latent factors.

Our goal in the simulation experiments was to determine the relationship between the observed latent factors $a, b, c, f$ and outcome risk $y$. We performed this in two ways. First, as a baseline, we regressed $\mathbf{y}$ on observed features $a, b, c, f$, to determine the approximate values of $a, b, c, f$ for each sample based on the relationship between $a, b, c, f$ and $\mathbf{a}, \mathbf{b}, \mathbf{c}, \mathbf{f}$ and $\mathbf{y}$. Then, assuming outcome risk $y$ was a second-order polynomial in $a, b, c, f$, we regressed the outcome risk $y$ on the coefficients determined in the previous regression, thereby estimating the functional relationship between $y$ and the latent factors $a, b, c, f$. Regression was performed using the Ridge model of scikit-learn package using three different values of the regularization parameter $\alpha$ [**?** ]. The initial regression coefficients corresponding to $\mathbf{a}$ through $\mathbf{f}$ were estimated on the larger dataset $\mathcal{D}_X$, then evaluated for the dataset $\mathcal{D}_Y$ for which outcome risk $y$ was available.

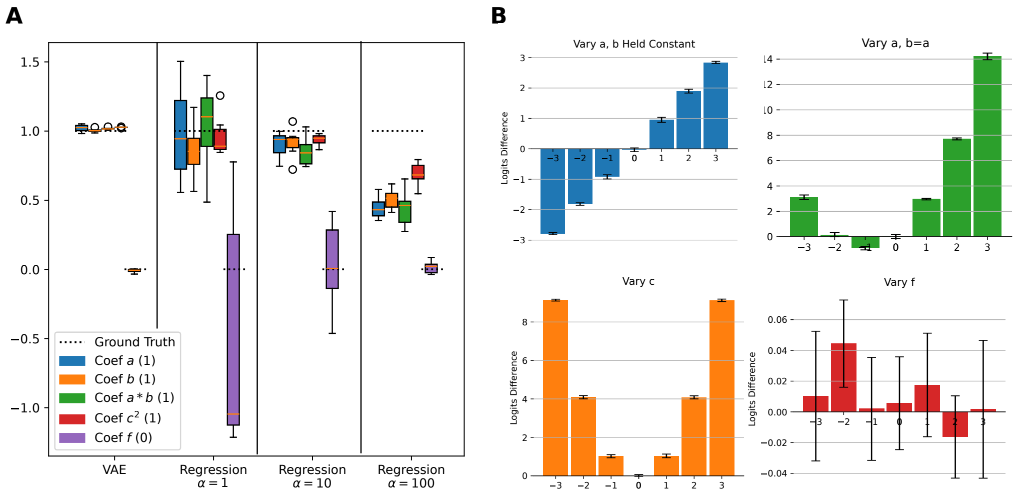


Figure 1: Results of the simulation experiments comparing use of the intervention VAE to two-stage regression in order to find the coefficients of a latent factor model of outcome risk. Part (A) shows estimates of coefficients for $a, b, a * b, c^2, f$ using either VAE or two-stage regression at three different settings for the regularization hyperparameter $\alpha$. It can be seen that the VAE is able to estimate coefficient values almost exactly while the two-stage regression has more variance and is highly dependent on the value of the regularization hyperparameter. (B) shows the method of estimating coefficients in the VAE model by intervening on each coefficient value and observing the resulting change in outcome risk.

| Coefficient MAE (True Value) | | | | | |
|---|---|---|---|---|---|
| | $a$ (1) | $b$ (1) | $a * b$ (1) | $c^2$ (1) | $f$ (0) |
| Reg. $\alpha = 1$ | $.106 \pm .092$ | $.052 \pm .060$ | $.095 \pm .085$ | $.019 \pm .020$ | $.906 \pm .472$ |
| Reg. $\alpha = 10$ | $.017 \pm .022$ | $.017 \pm .025$ | $.032 \pm .025$ | $.005 \pm .006$ | $.092 \pm .084$ |
| Reg. $\alpha = 100$ | $.314 \pm .079$ | $.247 \pm .065$ | $.332 \pm .138$ | $.101 \pm .051$ | $\mathbf{.001 \pm .002}$ |
| Inter. VAE | $\mathbf{.001 \pm .002}$ | $\mathbf{.001 \pm .002}$ | $\mathbf{.004 \pm .002}$ | $\mathbf{.001 \pm .001}$ | $\mathbf{.002 \pm .004}$ |

Table 9: Results of simulation experiments. Mean squared error for coefficients of latent factors leading to outcome risk determined using the intervention VAE or two-stage regression model. Three regression models are shown, using three different settings of the L2 regularization hyperparameter $\alpha$.
